# Parental Health Information Needs, Child-Health Literacy, and Use of Generative AI Among Parents of Children Aged 0 to 4 Years in France: A Cross-Sectional Study

**DOI:** 10.64898/2026.10.20.26365983

**Authors:** Kenza Gavaland, Nicolas Barascud, Anna-Karin Edstedt Bonamy, Pauline Martinot

**Affiliations:** Doctolib, Paris, France; Clinical Epidemiology Division, Department of Medicine, Solna, Karolinska Institutet, Stockholm, Sweden

**Keywords:** Information Seeking Behavior, Consumer Health Information, Digital Health, Pediatrics, Infant, Child, Preschool, Social Determinants of Health, Artificial Intelligence

## Abstract

**Background:** The first thousand days are a sensitive window for child health and social health inequalities. When routine care falls short, parents of young children turn to online sources and, recently, to generative artificial intelligence (AI). Evidence on their concerns, child-health literacy, and AI use is scarce. **Objective:** To describe health informational needs and concerns, child-health literacy, and use of AI for child-health questions, including differences by parental role and parity, among parents of young children. **Methods:** In this cross-sectional study, two anonymous self-administered online questionnaires were disseminated through a digital health application between May 12 and June 11, 2026, to French-speaking parents of at least one child aged 0 to 47 months in France. Questionnaire 1 covered unmet information needs, concerns across 16 topics, perceived intra-couple concordance, and confidence in routine care. Questionnaire 2 covered a four-item child-health literacy quiz and AI use (frequency, topics, 12-month trend, barriers, trust conditions, perceived effect on consultations, and disclosure). **Results:** In total, 12,634 parents responded (Questionnaire 1, n=5,410; Questionnaire 2, n=7,224), mostly mothers (94.6% and 92.9%). Most (61.8%) had searched for information without finding a clear, reliable answer, selecting a median of 4 (Interquartile range [IQR] 3-7) over 16 topics. Leading concerns were sleep (51.3%), nutrition (48.1%), and education (i.e., upbringing and behavior) (47.8%). Mothers were nearly twice as likely as fathers to select accident prevention (26.5% vs 16.3%) and parental mental health (38.7% vs 28.6%); first-time parents selected most topics more often than experienced parents, most markedly dental hygiene (43.8% vs 27.6%). The median child-health literacy score was 3 (IQR 3-4) on a 0-to-4 quiz, with 19.9% of parents scoring 2 or below; tooth-brushing age was the weakest item (68.4% correct). Mothers outscored fathers on every item. More than half of parents (54.2%) had used AI for a child-health question, 11.2% weekly; among those reporting a trend, 68.3% had increased AI use for child-health questions over the year and 6.2% decreased it. The main barriers to AI use were a preference for professionals (55.9%) and reliability concerns (48.7%); trust depended most on medical oversight of the tool (76.6%). AI use appeared additive, with 66.8% reporting no change in healthcare visit frequency. One third (35.5%) had told an AI something shared with neither a professional nor their network, rising to 64.1% among regular users. **Conclusions:** Parental informational needs were widespread and frequently unmet. AI has become a common and fast-rising source for child-health questions, at times a confidential one, used alongside rather than instead of professionals. Child-health literacy was generally high but uneven, with oral health the clearest gap. Strengths include the sample size and early-generative-AI timing; limitations include the cross-sectional design, self-report, digital-recruitment selection bias, and the small proportion of fathers.

## Introduction

The first years of a child’s life place heavy informational demands on parents. Decisions about feeding, sleep, vaccination, and everyday illness arise constantly, and they often exceed what routine care provides [1,2]. Concern about a young child’s development or behavior is common, yet these concerns often go unspoken in routine care, and fathers appear less well served than mothers [1–3]. When conventional care leaves these needs unmet, parents turn elsewhere: first to social media, where the quality of health content is uneven, and most recently to generative artificial intelligence, whose influence on parental behavior remains still poorly understood [4,5].

The first thousand days are a sensitive window for child health and for the social health inequalities that take root in this period, with effects that reach into adult life [6]. France treats this period as a public health priority: a national commission reported in 2020 and called for stronger institutional support and a digital health information platform for parents [7]. The information environment, however, has grown more difficult rather than easier, under the combined pressure of health misinformation on social media and the medicalization of ordinary infant behavior [4]. Whether a parent can find and judge reliable information in this setting depends heavily on health literacy.

Parental health literacy (i.e., the skills a parent needs to obtain and act on health information when raising a child) is an established determinant of child health [8]. It is often insufficient, especially in disadvantaged families, and low literacy predicts poorer child health and greater use of emergency care [9–11]. Self-reported health literacy may not fully reflect parents’ objectively assessed ability to understand and use health information [12], and basic misconceptions persist, at times among clinicians themselves [13]. Digital health information has become routine but uneven in quality, and in France its use varies with parental education level and age [14,15]. Evidence on generative AI is thinner: trust varies with a user’s profile [16] but does not track accuracy, since AI answers to medical questions are often unsupported by their own cited references [17], and parents have recently rated AI advice on child healthcare information as more trustworthy than a clinician’s [18].

### Knowledge gaps and objectives

Several questions remain open. Evidence on parental informational needs comes mostly from small convenience samples, and the most cited surveys predate the current digital ecosystem [1,2]. Few studies address the 0-to-4-year period specifically, when health decisions and preventive contacts are most frequent. Research on parental digital behavior largely predates conversational AI, leaving AI use in this population largely undocumented. Whether health literacy shapes how parents adopt and appraise AI-generated content is also unknown, a question with clear equity implications given the link between low literacy and social vulnerability [9,10], and one rarely examined in France despite the particular design of its preventive care [15]. To address these questions, a cross-sectional survey was implemented for parents of children aged 0 to 4 years in France. Parental health literacy level was expected to be linearly associated with the adoption of AI-based tools, and structural factors such as first-time parenthood and distance from care to be independently associated with heavier compensatory AI use. The survey had three aims: to characterize parental health concerns and informational needs in France in 2026, to describe health literacy and AI adoption across sociodemographic subgroups, and to document the prevalence and motivations of AI use in this population.

## Methods

### Study design

The INFANT study is a cross-sectional, observational survey of parents of young children in France, conducted through Doctolib, the country’s leading online health platform, used by more than 55 million people in France, over 81% of the national population, to manage their health [19]. Data were collected through a Typeform digital tool, with two self-administered questionnaires that were fielded separately and completed by different respondents: a first questionnaire on parental concerns and informational needs (Questionnaire 1) and a second on child-health literacy and the use of artificial intelligence tools (Questionnaire 2). Because both questionnaires were answered by distinct, anonymous samples, the dataset did not allow individual-level linkage between informational needs and AI use.

### Study population and eligibility

Respondents were eligible if they were the parent or legal guardian of at least one child aged 0 to 47 months, were aged 16 to 48 years, resided in France and were able to read and understand French. Individuals were excluded if they had no access to a computer, smartphone, or tablet, or could not read nor understand French well enough to complete the questionnaire.

### Recruitment and setting

Within the free Doctolib application, parents were invited through two placements: a marketing card, displayed to users whose Doctolib account recorded a dependent child under 4 years of age, and a drawer, a full-screen prompt shown when the app was opened. Tapping either redirected the parent to the questionnaire, hosted on Typeform. No financial incentive was offered, and responses were anonymous.

### Study period

Data collection took place in 2026 over two successive windows, one for each questionnaire: Questionnaire 1 was open from May 12 to May 22, and Questionnaire 2 from May 22 to June 11.

### Study instrument

Each questionnaire took about five minutes to complete. Both were designed from a review of the literature on parental health literacy, early childhood health, parental informational needs, and the use of AI tools in healthcare. No formal piloting or psychometric validation was carried out before release.

Both questionnaires began with the same three items on the child and respondent (the youngest child’s age, the child’s gender, and the respondent’s relationship to the child) and closed with the same optional, voluntary block on sociodemographic characteristics and lifestyle. Questionnaire 1 covered health informational needs and concerns for 0-to-4-year-old children, confidence in day-to-day parental care, and, for partnered respondents, a short block on perceived agreement with the partner; the couple items were shown only to respondents living with a partner. Questionnaire 2 covered child-health knowledge, assessed with a four-item quiz, and the use and perception of AI tools. A standing data-protection notice appeared on every free-text field, asking respondents not to enter identifying information and to describe only general themes.

Because the questionnaires used branching and matrix items, a single question count would be misleading, so the instrument is described by domain rather than by a fixed number of items. The full set of items, response options, and display conditions is provided in Multimedia Appendix 1, and a data dictionary listing every analyzed variable with its coding, derivation, source item, and the output in which it appears is provided as Multimedia Appendix 2.

### Outcomes and variables

The primary outcomes were parents’ health informational needs and their use of AI tools for child-health questions. Health informational needs were captured from a predefined list of sixteen health and parenting topics (item 1.1), from which each respondent could select any number of topics of interest and then name the single most difficult topic and the single current priority. AI use was measured as whether a parent had ever used an AI tool for a child-health question, together with how often (item 2.5).

Secondary outcomes were confidence in day-to-day care, child-health literacy, perceived agreement with the partner among coupled respondents, and parents’ wider relationship with AI tools, covering query topics, the twelve-month trend in use, situations judged useful, barriers to use, conditions for trust, the perceived effect on professional consultations, and disclosure to AI of concerns not shared with a clinician or social network. Two summary scores were derived: a care-task confidence score as the mean of five Likert items (item 1.7), and a child-health literacy score as the number of the four quiz items answered correctly (items 2.1 to 2.4). Perceived within-couple concordance was derived by comparing each respondent’s priority topic with their estimate of the partner’s priority (items 1.3 and 1.4). Sociodemographic and lifestyle characteristics were treated as descriptive variables and as candidate predictors of AI use. The lifestyle block and the two free-text fields were collected but not included in the present analyses. Full definitions and coding appear in Multimedia Appendix 2.

### Statistical analysis

All analyses were performed in Python 3.12, using SciPy (v1.17.1), statsmodels (v0.14.6), and scikit-learn (v1.8.0) for statistical computation, pandas (v3.0.3) and NumPy (v2.4.6) for data handling, and Matplotlib (v3.10.9) and seaborn (v0.13.2) for figures. The analysis code was organized as a versioned package, and every stochastic procedure was run under a fixed random seed (42) for reproducibility. Both questionnaires were completed by independent respondent cohorts with no linking identifier (Questionnaire 1, n = 5,410; Questionnaire 2, n = 7,224), so every analysis was reported within a single questionnaire and no cross-questionnaire record linkage was attempted.

Categorical variables were summarized as counts and percentages, computed within each questionnaire, and continuous variables as mean (standard deviation, SD) or median (interquartile range, IQR) as appropriate. Responses of “prefer not to say” or “don’t know” were recoded as missing throughout. Multi-select blocks (i.e., twelve-month trend in AI use, useful situations, barriers, trust prerequisites, and disclosure topics) were decomposed into one binary indicator per option.

Subgroup comparisons of categorical variables used Pearson’s chi-square test of independence, with Fisher’s exact test substituted for 2×2 tables whenever the minimum expected cell count fell below 5. Continuous outcomes were compared between two groups with the two-sided Mann-Whitney U test, and across more than two groups with the Kruskal-Wallis test. All tests were two-sided, with α=.05. To control the false discovery rate across the multiple subgroup tests within each table (e.g., the sixteen concern domains), p-values were adjusted with the Benjamini-Hochberg procedure within each family of tests; both raw p-values and adjusted p-values were reported.

Unless stated otherwise, the 95% confidence intervals shown in figures and supplementary tables were obtained by nonparametric bootstrap (i.e., 2,000 resamples, percentile method) on the statistic displayed, whether a proportion, a mean, or an odds ratio, giving a single, internally consistent interval-estimation procedure throughout the paper.

As a sensitivity analysis for the parent-role differences in concern prevalence, per-topic logistic regressions of each concern indicator were fitted on the parental role (father as reference), reporting odds ratios with bootstrap 95% confidence intervals and adjusted p-values over the same sixteen-test family. To account for within-respondent correlation across topics, a single binomial generalized estimating equation (GEE) model was fitted with an exchangeable working correlation, clustered by respondent, and the joint topic-by-role interaction was tested with a Wald omnibus test.

The child-health literacy score was computed as the sum of four binary knowledge items (i.e., recommended sleep position, age to begin oral hygiene, age to begin complementary feeding, and being aware of a developmental warning sign), each scored 0 or 1, for a score of 0 to 4.

Predictors of any AI use for child-health questions were examined by logistic regression. Unadjusted odds ratios came from univariate complete-case models. Adjusted odds ratios came from a multivariable model including first-time parenthood, child age band, parent age (per 5 years), literacy score, area of residence, ease of access to a healthcare professional, educational attainment, chronic condition in the child, and parent role.

The minimum sample size for estimating a single proportion was calculated from the formula n = Z²p(1 − p)/d², where p is the expected proportion. Assuming a 95% confidence level (Z = 1.96), an anticipated proportion of 50% (the most conservative value), and a margin of error of 3% (d = 0.03), this gave approximately 1,068 participants per questionnaire.

The achieved samples far exceeded this target, with 5,410 respondents to the concerns questionnaire (Questionnaire 1) and 7,224 to the literacy-and-AI questionnaire (Questionnaire 2), so the realized precision was high, with a margin of error of about ±1.3% or less per questionnaire at 50% prevalence and 95% confidence.

### Missing data

The multivariable model drew on nine predictors: three binary (first-time parenthood, chronic condition in the child, and parent role), five ordinal (child age band, literacy score, area of residence coded by population size of the place of residence, ease of access to a healthcare professional, and educational attainment), and one continuous (parent age). Three were complete, since child age band, parent role, and the derived literacy score came from mandatory items. The remaining six belonged to the optional sociodemographic block and carried missingness of about 28% to 45%, highest for first-time parenthood and parent age. Rather than restrict the model to complete cases, missing predictor values were filled by multiple imputation by chained equations (MICE) (20 imputations, 10 burn-in iterations), and estimates were pooled across the imputed datasets by Rubin’s rules, with 95% Wald confidence intervals. The fraction of missing information ranged from 0.03 to 0.64 and exceeded 0.40 for first-time parenthood and parent age, whose adjusted estimates should be read with more caution. Multimedia Appendix 4 reports the unadjusted odds ratios from univariate complete-case models alongside the adjusted odds ratios from the imputed model. Employment status was left out of the model, being a five-level nominal variable with about 29% missingness; its unadjusted association with AI use is reported in Multimedia Appendix 5.

Perceived intra-couple concordance was assessed with Cohen’s κ between each respondent’s own top-priority concern and their reported expectation of their partner’s top priority. Because the survey collected one response per respondent rather than paired dyadic data, this κ quantifies within-respondent perceived agreement rather than actual couple concordance, and is interpreted accordingly.

### Ethics and data protection

The study was conducted in accordance with the Declaration of Helsinki. As an observational questionnaire survey involving no personal data, no intervention, no biological sampling, and no reuse of individual medical records, the study did not fall within the scope of the Regulation (EU) 2016/679 of 27 April 2016 on the protection of natural persons with regard to the processing of personal data and on the free movement of such data (General Data Protection Regulation, GDPR), or the French Data Protection Act (Loi n° 78-17 du 6 janvier 1978 relative à l’informatique, aux fichiers et aux libertés), or, where applicable, the French Public Health Code and specifically the French framework for research involving the human person (Loi n° 2012-300 du 5 mars 2012 relative aux recherches impliquant la personne humaine, known as the Loi Jardé). Approval by a Committee for the Protection of Persons (Comité de Protection des Personnes) was therefore not required.

Before starting, participants were shown a short notice describing the purpose of the questionnaire, stating that responses were anonymous, and linking to Doctolib’s personal-data protection policy regarding the possibility to push forth voluntary questionnaires under Doctolib’s legitimate interest. Participation in the study was entirely voluntary, and participants were free to withdraw at any time. The questionnaire was designed to collect only anonymous, non-identifying information, such as general responses, age bands, and broad categories for other variables. No personal data was requested. Each free-text field included a reminder asking participants not to enter information that could identify them directly or indirectly. Therefore the responses themselves did not constitute personal data within the meaning of the GDPR. Responses were exported from Typeform to secure, French health-data-hosting-certified (Hébergeur de Données de Santé, HDS) Doctolib servers, with access restricted to the research team, and all analyses were performed on aggregated data.

## Results

### Population characteristics

The two samples were sociodemographically comparable (Table 1). Mothers made up the large majority of both (94.6% in Questionnaire 1, 92.9% in Questionnaire 2), fathers 5.4% and 7.1% respectively. The child gender distribution was balanced (51.1% girls in Questionnaire 1, 49.7% in Questionnaire 2). Children spanned the full range to 47 months, the 3-to-4-year band being the most frequent (28.4% in Questionnaire 1, 31.9% in Questionnaire 2), with a median child age of 18.0 months (IQR 9.5-42.0) in Questionnaire 1 and 30.0 (IQR 9.5-42.0) in Questionnaire 2.

**Table 1.** Sociodemographic and health-related characteristics of respondents, by questionnaire.^a,b,c^.

| Characteristic | Questionnaire 1 (n=5,410) | Questionnaire 2 (n=7,224) |
| --- | --- | --- |
| **Parent role** |  |  |
| Mother | 5,116 (94.6) | 6,709 (92.9) |
| Father | 294 (5.4) | 515 (7.1) |
| **Child gender** |  |  |
| Female | 2,764 (51.1) | 3,593 (49.7) |
| Male | 2,604 (48.1) | 3,579 (49.5) |
| Missing | 42 (0.8) | 52 (0.7) |
| **Child age band** |  |  |
| Under 3 months | 697 (12.9) | 678 (9.4) |
| 3-6 months | 584 (10.8) | 672 (9.3) |
| 7-12 months | 506 (9.4) | 588 (8.1) |
| 1-2 years | 1,028 (19.0) | 1,453 (20.1) |
| 2-3 years | 1,061 (19.6) | 1,527 (21.1) |
| 3-4 years | 1,534 (28.4) | 2,306 (31.9) |
| **Family status<sup>b</sup>** |  |  |
| Single parent | 866 (16.0) | — |
| Partnered | 4,544 (84.0) | — |
| **First-time parent** |  |  |
| No | 2,128 (39.3) | 2,044 (28.3) |
| Yes | 2,164 (40.0) | 1,962 (27.2) |
| Missing | 1,118 (20.7) | 3,218 (44.5) |
| **Total number of children at home** |  |  |
| 1 | 2,100 (38.8) | 2,364 (32.7) |
| 2 | 1,486 (27.5) | 1,868 (25.9) |
| 3 | 607 (11.2) | 714 (9.9) |
| 4 | 196 (3.6) | 223 (3.1) |
| 5 | 58 (1.1) | 78 (1.1) |
| 6 | 45 (0.8) | 43 (0.6) |
| Missing | 918 (17.0) | 1,934 (26.8) |
| **Highest diploma** |  |  |
| No diploma | 223 (4.1) | 221 (3.1) |
| Brevet, CAP, or BEP | 663 (12.3) | 745 (10.3) |
| High-school graduation | 976 (18.0) | 1,084 (15.0) |
| Bac+2 (BTS, DUT) | 664 (12.3) | 701 (9.7) |
| Bachelor's degree (Bac+3) | 771 (14.3) | 883 (12.2) |
| Master's degree (Bac+5) | 975 (18.0) | 1,081 (15.0) |
| Doctorate (Bac+8) | 110 (2.0) | 125 (1.7) |
| Missing | 1,028 (19.0) | 2,384 (33.0) |
| **Employment status** |  |  |
| Full-time employed | 1,942 (35.9) | 2,506 (34.7) |
| Part-time employed | 686 (12.7) | 804 (11.1) |
| Maternity/paternity/parental leave | 764 (14.1) | 791 (10.9) |
| Unemployed or job-seeking | 954 (17.6) | 986 (13.6) |
| Student | 52 (1.0) | 39 (0.5) |
| Missing | 1,012 (18.7) | 2,098 (29.0) |
| **Area of residence** |  |  |
| Large city (>100,000) | 1,220 (22.6) | 1,301 (18.0) |
| Mid-size town (2,000-100,000) | 2,109 (39.0) | 2,465 (34.1) |
| Rural or village (<2,000) | 1,072 (19.8) | 1,414 (19.6) |
| Missing | 1,009 (18.7) | 2,044 (28.3) |
| **Parent age band** |  |  |
| Under 25 | 459 (8.5) | 355 (4.9) |
| 25-29 | 630 (11.6) | 815 (11.3) |
| 30-34 | 1,445 (26.7) | 1,601 (22.2) |
| 35-39 | 1,291 (23.9) | 1,343 (18.6) |
| 40-44 | 425 (7.9) | 347 (4.8) |
| 45+ | 63 (1.2) | 50 (0.7) |
| Missing | 1,097 (20.3) | 2,713 (37.6) |
| **Ease of access to a health professional** |  |  |
| Very difficult | 633 (11.7) | 720 (10.0) |
| Fairly difficult | 1,071 (19.8) | 1,233 (17.1) |
| Fairly easy | 1,374 (25.4) | 1,539 (21.3) |
| Very easy | 1,336 (24.7) | 1,521 (21.1) |
| Missing | 996 (18.4) | 2,211 (30.6) |
| **Child chronic condition** |  |  |
| No | 3,260 (60.3) | 3,936 (54.5) |
| Yes | 925 (17.1) | 928 (12.8) |
| Missing | 1,225 (22.6) | 2,360 (32.7) |

- Categorical variables are n (%); percentages are column percentages and may not total 100% because of rounding and, where shown, missing responses. - Family status was collected in Questionnaire 1 only; — indicates not collected. - Parent age was available for 4,313 respondents in Questionnaire 1 and 4,511 in Questionnaire 2. Age was collected as a banded birth year in Questionnaire 1 and a banded current age in Questionnaire 2; to report it on a single scale in this table, each band was converted to its midpoint and the two sets of bands harmonized into the six age bands shown here

In Questionnaire 1, 84.0% of respondents lived with a partner and 16.0% were single parents. First-time and experienced parents were present in similar proportions among those who answered the item (40.0% and 39.3% in Questionnaire 1; 27.2% and 28.3% in Questionnaire 2). Educational attainment spanned all levels, with about one in five holding a master’s degree or higher, and residence was distributed across mid-size towns (39.0%), large cities (22.6%), and rural areas (19.8%). Reported ease of access to a health professional ranged from very difficult (11.7%) to very easy (24.7%), and 17.1% of Questionnaire 1 respondents reported a chronic health condition in their child. Missing data were more frequent on the optional sociodemographic block, particularly in Questionnaire 2 (for example, 37.6% missing for the parent age band).

### Parental informational needs and concerns

In Questionnaire 1, 3,344 parents (61.8%) reported having looked for health or parenting information without finding a clear and reliable answer. Parents selected a median of 4 (IQR 3-7) topics of interest from the sixteen-item list. The most frequently selected topics were sleep (51.3%), feeding and nutrition (48.1%), upbringing and behavior (47.8%), self-confidence and emotions (44.9%), and common childhood illnesses (39.5%), while the least selected were vaccination (23.6%), childcare (14.7%), and safety and transport (12.5%).

Mothers and fathers differed in their topic profile (Table 2; GEE topic-by-role interaction P<.001). After adjustment for multiple testing, mothers were more likely than fathers to select accident prevention (OR 1.85, 95% CI 1.38-2.59), upbringing and behavior (OR 1.67, 1.31-2.12), parental mental health (OR 1.58, 1.24-2.06), common illnesses (OR 1.54, 1.21-1.98), and self-confidence and emotions (OR 1.43, 1.11-1.81); the remaining topics did not differ significantly by parental role (Multimedia Appendix 7; the same per-topic odds ratios are shown as a forest plot in Multimedia Appendix 8). First-time parents selected more topics than experienced parents on most items, with significant differences for sleep, feeding, common illnesses, dental hygiene, work-family balance, accident prevention, screens, motor development, childcare, and safety and transport (all adjusted P<.05), each in the direction of higher selection by first-time parents (Table 2).

**Table 2.**
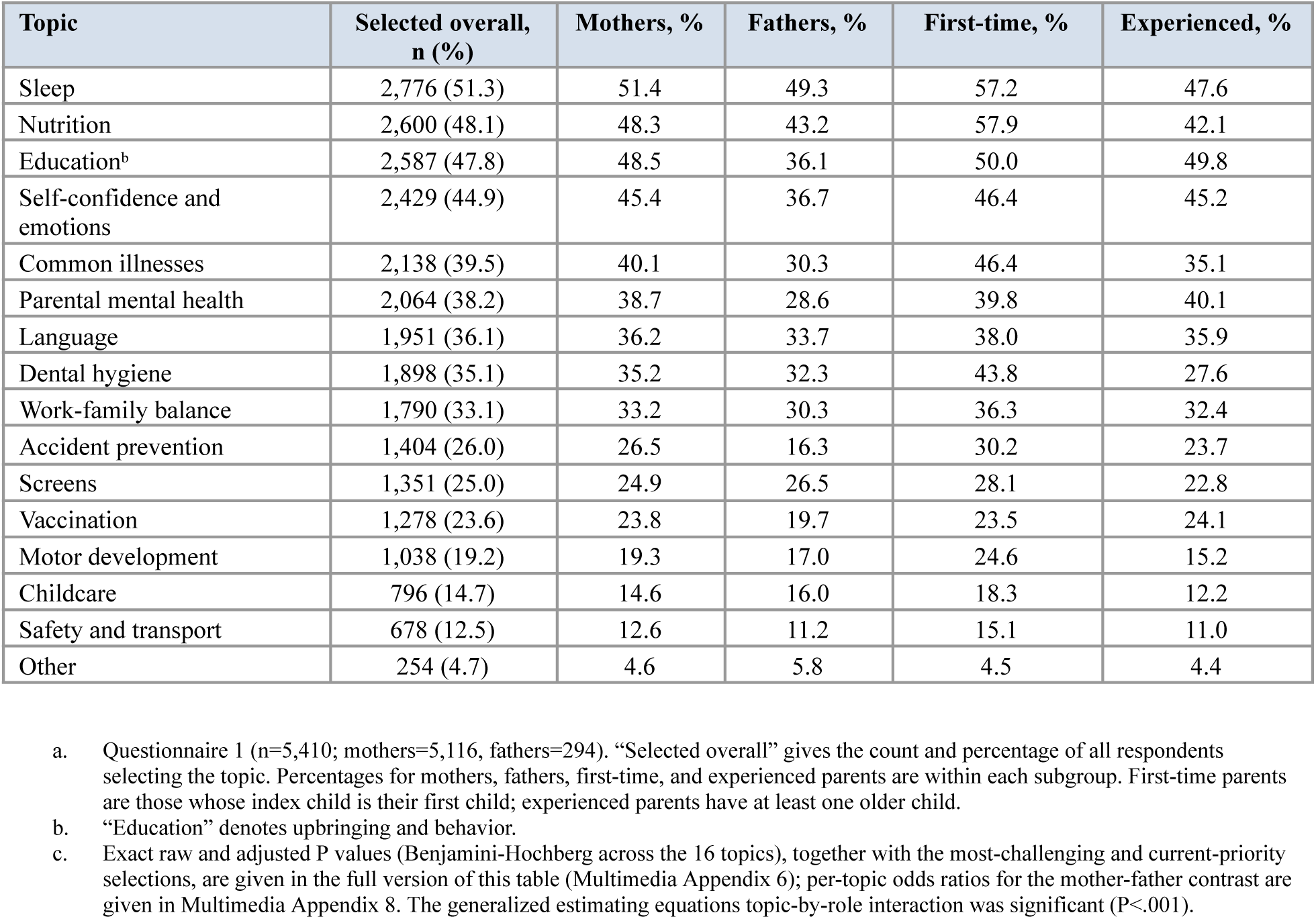
Informational needs and concerns among parents, by parental role and parity (abridged).^a,b,c^.

### Perceived intra-couple concordance

Among coupled respondents (n = 4,544), 45.1% perceived their priority topic to coincide with their partner’s, with agreement between the respondent’s priority and the priority attributed to the partner (Cohen κ=.368) (Multimedia Appendix 9). Fathers perceived agreement more often than mothers (58.7% vs 44.3%, adjusted *P*<.001). About one quarter of respondents (26.5%) reported that their concerns diverged from their partner’s, more frequently mothers than fathers (27.3% vs 15.7%, adjusted *P*<.001). Perceived divergence was closely tied to differing priorities: among respondents whose stated priority differed from the one they attributed to their partner, 42.3% reported diverging concerns, compared with 8.8% among those whose priorities coincided (*P*<.001). The topics most often named as sources of disagreement were upbringing (19.6%), sleep (14.9%), work-family balance (11.4%), feeding (10.4%), and screens (10.3%). These figures reflected the respondent’s perception of the partner’s priority rather than the partner’s own answer, as couples were not surveyed as dyads.

### Parental confidence in daily care

Confidence in managing the child’s day-to-day health was high across all items (Multimedia Appendix 10, Multimedia Appendix 11). Item means ranged from 4.12 (SD 0.84) for general health questions and 4.26 (SD 1.05) for cleaning the child’s nose to 4.92 (SD 0.39) for changing the diaper, on a 1-to-5 scale; the composite confidence score averaged 4.62 (SD 0.51). Mothers and fathers differed significantly on two tasks after adjustment, changing the diaper (4.92 vs 4.79, adjusted *P*<.001) and bathing and trimming nails (4.69 vs 4.45, adjusted *P*<.001), while the composite did not differ significantly by role. Experienced parents were more confident than first-time parents on every individual task except changing the diaper, as well as on the composite score, the mean of the five care-task items (4.69 vs 4.55, adjusted *P*<.001). The characteristics most strongly associated with confidence were educational attainment and ease of access to a health professional: higher education and easier access each predicted greater confidence on almost every task and on the composite score (Multimedia Appendix 11).

### Child-health literacy

In Questionnaire 2, the composite literacy score had a median of 3 (IQR 3-4) of 4, and about one parent in five (19.9%) scored 2 or below. The proportion answering correctly was highest for infant sleep position (89.3%) and the walking warning sign at 2 years (83.2%), intermediate for the timing of complementary feeding (78.4%), and lowest for the age to begin tooth-brushing (68.4%) (Multimedia Appendix 12). Mothers scored higher than fathers on every item and on the composite (3.24 vs 2.60, *P*<.001), with the largest gap on tooth-brushing age (69.8% vs 50.3% correct). Parity was not associated with the composite score (first-time 3.26 vs experienced 3.24, *P*=.452) or with any individual item. The composite score was strongly associated with educational level and parental age (both adjusted *P*<.001) and, to a lesser degree, with ease of access to a healthcare professional (adjusted *P*=.004), but not with residence or a chronic condition in the child (Multimedia Appendix 13).

### Use of AI for child-health questions

More than half of Questionnaire 2 respondents (n = 3,919, 54.2%) reported having used a conversational AI tool for a child-health question (Figure 1). Use was occasional for most: 30.2% had used it once or twice out of curiosity, 12.8% several times per month, and 11.2% several times per week. Among the 3,919 users, the most common query topics were symptoms and illnesses (59.9%), activity and play ideas (47.6%), sleep (42.0%), feeding (38.9%), behavior and emotions (36.6%), child development (35.8%), and everyday practical questions (35.3%); vaccination was the least common (11.5%). Among the 3,089 respondents who could report a direction of change over the preceding 12 months, use had increased for 68.3% (37.4% greatly, 30.9% slightly) and decreased for 6.2%, with 25.5% reporting no change (Figure 1). When asked in which situations an AI assistant would be useful, respondents most often cited everyday questions not requiring a doctor (44.9%), reassurance about minor worries (38.3%), and nighttime triage (29.6%); 25.2% felt no situation would be useful, and parents selected a median of 1 (IQR 1-3) useful situation (Multimedia Appendix 14). These perceptions differed sharply by use: AI users endorsed every situation far more often than non-users, while non-users predominated in seeing no useful situation at all (47.4% vs 6.4%) (Multimedia Appendix 15).

**Figure 1.**
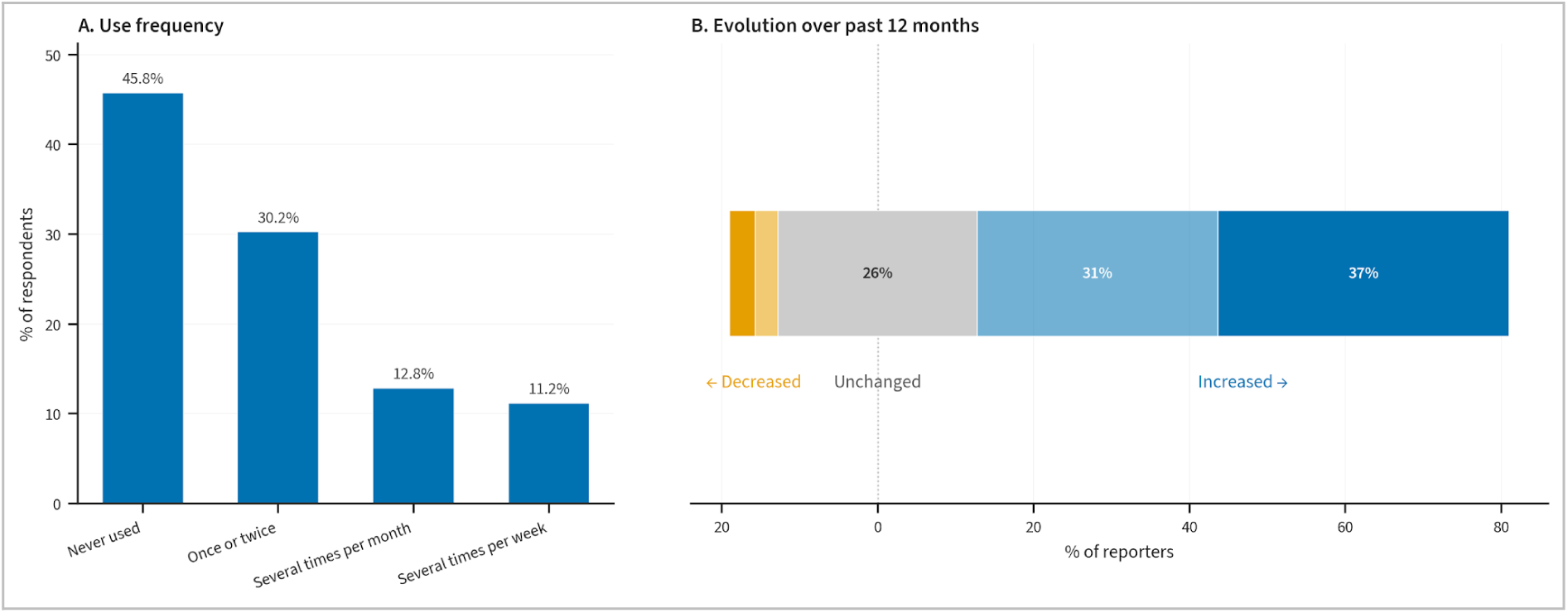
Patterns of conversational artificial intelligence (AI) use for child-health questions among parents (Questionnaire 2). (A) Frequency of AI use among all respondents (n=7,224); bars show the percentage of respondents in each category, from never used to several times per week. (B) Self-reported change in the frequency of AI use over the preceding 12 months, among the 3,089 respondents who could report a direction of change (reporters; parents not using AI one year earlier are excluded). The diverging stacked bar shows the percentage of reporters whose use decreased (left, orange), remained unchanged (center, gray), or increased (right, blue), with the increase divided into slightly and greatly. AI: artificial intelligence.

### Predictors of AI use

Several characteristics were independently associated with AI use (Figure 2; Multimedia Appendix 4). The adjusted odds of using AI were higher for first-time parents (adjusted OR 1.47, 95% CI 1.27-1.70) and rose with a higher literacy score (1.17, 1.11-1.24 per additional correct answer) and with more urban residence (1.15, 1.07-1.25 per level). The adjusted odds were lower for older children (0.88, 0.86-0.91 per age band), older parents (0.89, 0.83-0.94 per 5 years), and easier access to a health professional (0.92, 0.86-0.97 per level). Parental role, educational level, and a chronic condition in the child were not associated with AI use after adjustment. Unadjusted subgroup rates were consistent with these results (Multimedia Appendix 5): AI use was higher among first-time parents (60.1% vs 49.5%), declined with child age (from 65.6% under 3 months to 47.9% at 3 to 4 years) and with parental age (about 66% among parents in their twenties, falling to 42.7% at 42 to 47 years), and rose with the literacy score, while it did not differ significantly by parental role, education, ease of access to care, or a chronic condition.

**Figure 2.**
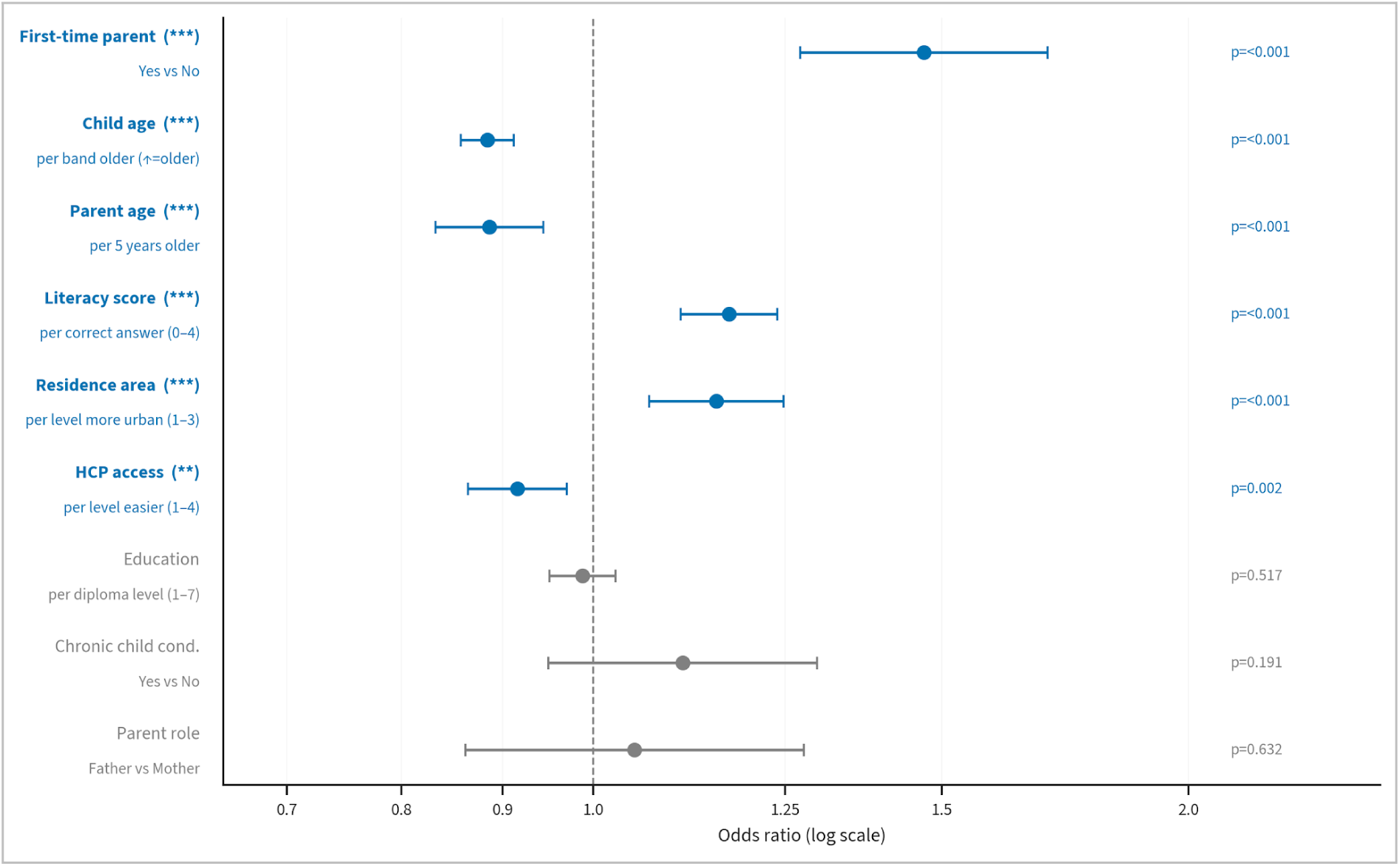
Independent predictors of conversational artificial intelligence (AI) use for child-health questions (Questionnaire 2; n=7,224). Points are adjusted odds ratios and horizontal bars are 95% confidence intervals from a multivariable logistic regression pooled across 20 datasets imputed by multiple imputation by chained equations (MICE) with Rubin’s rules. The x-axis is on a logarithmic scale; the dashed line marks an odds ratio of 1 (no association), and values above 1 indicate higher odds of AI use. Each predictor is modeled per the unit shown on the axis. Education, chronic child condition, and parental role were not significant. Significance is from the Wald test in the pooled model (*P<.05, **P<.01, ***P<.001). The fraction of missing information for each predictor is reported in Multimedia Appendix 4. HCP: health care professional; MICE: multiple imputation by chained equations.

### Barriers, trust, perceived impact, and disclosure

The most common reasons for not using AI more were a preference for talking to a professional (55.9%) and concerns about reliability (48.7%); privacy concerns (14.5%) and discomfort with the tools (6.6%) were less frequent (Figure 3; Multimedia Appendix 16). These barriers varied steeply across user categories: a preference for professionals fell from 64.9% among non-users to 38.2% among regular users, while “nothing holds me back” rose from 8.3% to 41.0% (both adjusted *P*<.001). The conditions most often required to trust an AI tool were supervision by physicians (76.6%) and certification by an official authority (57.9%), both endorsed more by users than non-users (physician supervision rising from 71.0% to 82.4% across categories, adjusted *P*<.001); a recommendation from one’s own doctor (34.2%) shifted little across categories. Among the 3,689 AI users who answered, most reported no change in how often they consulted a health professional (66.8%) (Multimedia Appendix 16); 18.9% said they came to consultations better prepared, 6.9% consulted less often, and 7.4% more often. One third of all Questionnaire 2 respondents (35.5%) reported having raised something with an AI tool that they had shared with neither a health professional nor their personal network, most often concerning their own well-being (25.2%) or doubts about their parenting role (10.9%). Disclosure rose with intensity of use, from 15.4% of non-users to 43.2% of occasional users and 64.1% of regular users; among regular users, 50.9% reported disclosing well-being concerns and 26.8% parenting doubts.

**Figure 3.**
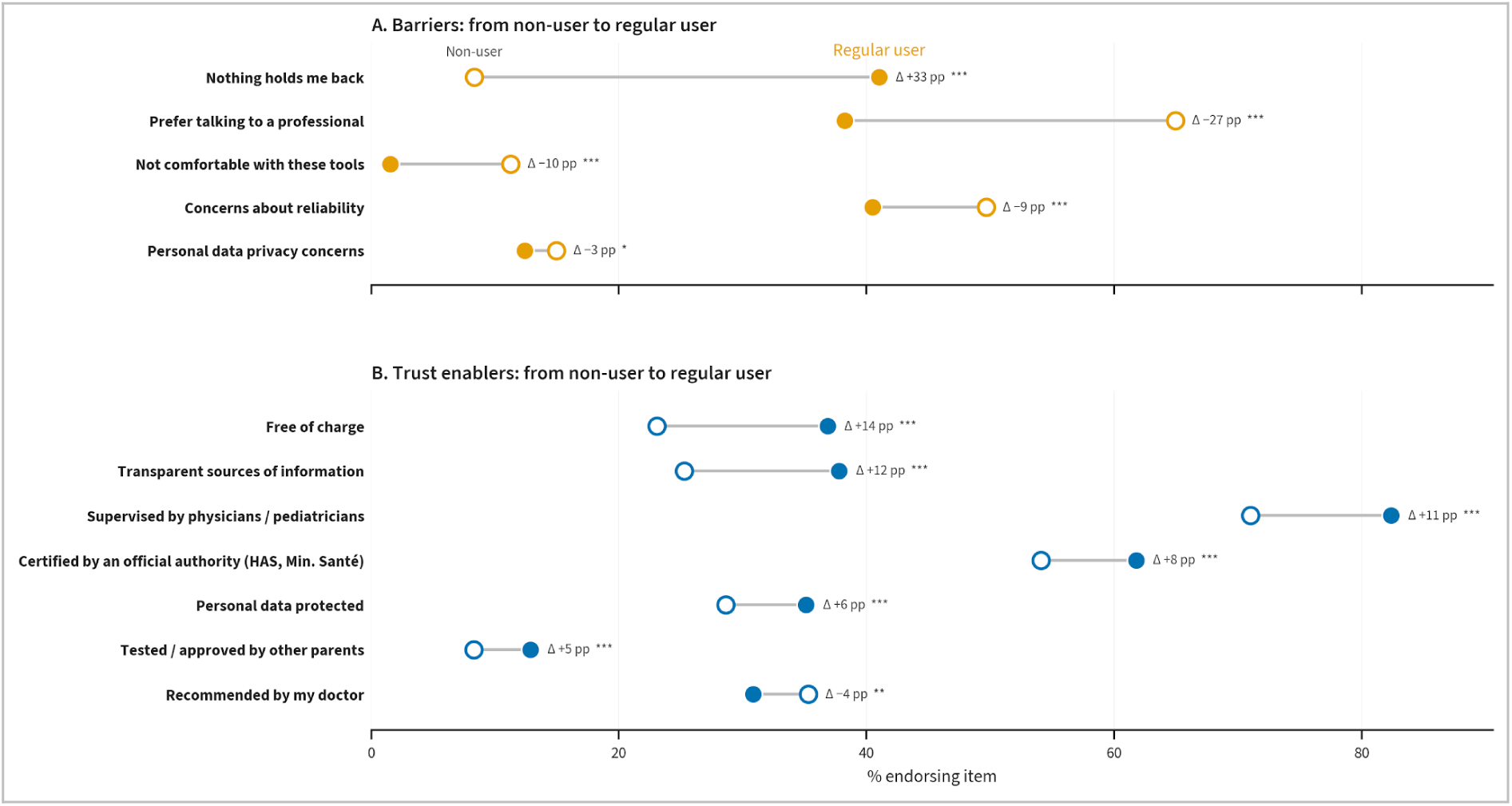
Perceived barriers to and trust enablers for conversational artificial intelligence (AI) use, from non-users to regular users (Questionnaire 2; non-users n=3,305, regular users n=1,734). (A) Barriers; (B) trust enablers. Each row connects the percentage of non-users (open circle) and regular users (filled circle) endorsing the item, and the annotation gives the change in percentage points between the two groups. The x-axis is the percentage of each group endorsing the item; error bars are bootstrap 95% confidence intervals. Significance is from a chi-square test across the three user categories (non-users, occasional users, and regular users), adjusted with the Benjamini-Hochberg procedure (*P<.05, **P<.01, ***P<.001).

## Discussion

### Principal Findings

This study describes what parents of 0-to-4-year-old children in France currently want to know about their child’s health in 2026, how confident they feel handling it, what they actually know, and how they are starting to use generative AI to reduce their health knowledge gaps. Among more than 5,400 respondents to the concerns questionnaire, 61.8% had searched for health or parenting information without finding a clear and reliable answer, and parents selected a median of 4 (IQR 3-7) of 16 topics, with concerns concentrated on sleep (51.3%), feeding (48.1%), upbringing and behavior (47.8%), self-confidence and emotions (44.9%), and common childhood illnesses (39.5%). Confidence in routine care was uniformly high (item means 4.12 to 4.92 on a 5-point scale) but tracked education and ease of access to a clinician more than gender or parity. Knowledge was generally sound (literacy median 3, IQR 3-4, of 4) yet incomplete: about one parent in five scored 2 or below, and 32% did not know that tooth-brushing should begin with the first tooth. More than half of the parents in the AI questionnaire (54.2%) had already used a conversational tool for a child-health question, most often about symptoms (59.9%), and reported use was rising steeply over the preceding year. For most users AI complemented professional care rather than replacing it, and a third of parents had raised something with an AI tool that they had told neither a clinician nor anyone in their circle. Across the sample, trust depended far more on professional oversight (76.6%) and official certification (57.9%) than on cost or peer approval.

### Comparison With Prior Work

The level of unmet informational need fits a long line of evidence that concern about young children is common and often goes unaddressed in routine care. Our results are in line with a Dutch population sample study, where 49.3% of parents reported concern about at least one aspect of their child’s development or behavior, with higher rates among parents of the youngest children and among socially vulnerable families, reaching 60% below the poverty line [1]. A decade earlier, a US survey found that only 28% of mothers raised non-medical concerns with their child’s pediatrician [2]. The 61.8% of parents here who could not find a clear answer sits at the upper end of that range and suggests that the distance between what parents want to know and what the standard care pathway provides has not narrowed, even as information has become easier to obtain. The order of concerns, with sleep, feeding, upbringing, and emotional questions ranking above acute illness, matched earlier work showing that everyday developmental and behavioral questions, rather than medical emergencies, dominated parents’ needs [1,20]. This environment was also shaped by recent health misinformation on social media and by the medicalization of normal infant behavior, which raise the stakes of where parents turn for answers [4].

Differences between mothers and fathers ran in the same direction across both instruments. Mothers were more likely than fathers to flag several topics, notably accident prevention, upbringing and behavior, parental mental health, common illnesses, and self-confidence and emotions, and their overall profile of concerns differed by parental role. Mothers also outscored fathers on every literacy item, most markedly on tooth-brushing age (69.8% vs 50.3% correct). These patterns are consistent with reports that fathers are less engaged with, and less well served by, child-health information and support, including a Swedish study in which fathers described difficulty managing the demands of early parenthood [3]. These results have to be read against the heavily maternal makeup of the sample, a bias considered below.

A gap between confidence and knowledge appeared once both genders were compared. Fathers rated their confidence with general health questions on par with mothers, yet scored well below them on the quiz, a mismatch consistent with evidence that self-assessed health confidence may not reflect objectively assessed knowledge [12]. The same caution applies to the high overall confidence in routine care; ceiling-level self-ratings should not be mistaken for adequate health literacy, since literacy may be overstated when inferred from confidence alone [21]. More informative than gender or parity was the social patterning of both confidence and knowledge. Higher educational attainment and easier access to a clinician were each associated with greater confidence on most care tasks, and education was strongly associated with the literacy composite. The direction was in line with a large German survey (n = 4 217) in which 45.8% of parents had problematic or inadequate health literacy, with socioeconomic position the strongest predictor and low literacy linked to poorer nutritional and dental-care practices and reduced preventive care [9], and given systematic-review evidence connecting low parental health literacy to worse child outcomes and higher emergency-department use [10,11]. That the same structural factors shaped both perceived ease and measured knowledge here points to a disparity that operates regardless of who the parent is, and that deserves attention when parent-facing resources are designed [8].

Experience showed a split effect that is itself informative. Experienced parents were more confident than first-time parents on every care task, and first-time parents selected more informational needs across almost every topic, consistent with an information demand that peaks with a first child. Parity made no difference to quiz performance, however. Knowing when to introduce solids or to start brushing is learned from a source rather than acquired by repetition, so having raised a child did not translate into better factual knowledge. Experience appears to reassure parents without correcting specific gaps, of which the tooth-brushing item (32% incorrect) is the clearest and most actionable for preventive messaging.

More than half of the parents had used generative AI for a child-health question. That is lower than the near-universal use of general digital sources reported before conversational tools appeared, such as the 91% of Swiss parents who used the internet for child-health information, most of whom doubted what they found and two-thirds of whom wanted their pediatrician to point them to trustworthy sources [14]. This level is nonetheless high for so recent a technology, and use is rising quickly: among parents who could report a direction of change, increases over the past year outnumbered decreases by roughly eleven to one (68.3% vs 6.2%). How parents used AI followed the medium-by-concern pattern described before, in which the internet served developmental and lifestyle questions while professionals stayed the preferred channel for acute or sensitive matters [22]. Symptoms and everyday questions led the AI queries, while vaccination was the least common topic (11.5%), far below its weight as a general concern. In France, vaccine attitudes are closely tied to health literacy and to confidence in the health-care system [23], consistent with an interpretation in which parents keep vaccination anchored to institutional and professional sources rather than treating it as an AI question. The gradients seen in confidence and knowledge are likewise consistent with French evidence that online health-information use is unequally distributed by education, age, and socioeconomic position [15], and with survey work showing that trust in, and willingness to act on, AI health advice varies with user profile and prior knowledge [16,24].

The characteristics that predicted AI use pointed towards an AI adoption motivated by health information need and digital engagement rather than by social disadvantage nor limited resources. Using a multivariable model, parents were more likely to use AI if they had a first child, a younger child, or were younger themselves, in line with the higher information demand of early and first-time parenthood that the needs data also showed. AI adoption was likewise higher among parents with greater health literacy and among urban residents, and lower among those reporting easier access to a health professional. The literacy result ran against the study’s initial expectation that lower literacy would drive compensatory AI use: more literate parents used AI more, not less, which fits French evidence that online health-information use is concentrated among more educated and information-engaged groups [15]. Easier access to a clinician went with slightly lower use, which gives only weak support to the idea that AI compensates for poor professional access. The geographic finding cuts the other way: if AI were filling the gap left by distance from services, AI use should have been significantly highest among rural parents, yet it rose with urban residence instead. Educational level, parental role, and a chronic condition in the child were not independently associated with use. Because adoption is concentrated among more literate and urban parents, an AI channel left to diffuse on its own could widen rather than narrow existing information inequalities. These associations concern whether parents adopt AI, not how critically they appraise what it returns, which this study did not measure.

The conditions parents set for trusting an AI tool were clear and consistent. Roughly 75% would trust such a tool if physicians supervised it, and a majority wanted certification by an official authority, while peer approval and free access counted for much less. Reliability concerns (48.7%) and a preference for speaking with a professional (55.9%) were the leading barriers, and privacy concern was comparatively low (14.5%). This retained caution is reassuring against the documented risk of excessive trust, since a large share of large-language-model answers to medical questions are not fully supported by the sources they cite [17], and parents have rated AI-generated pediatric advice as more trustworthy and accurate than clinician-written advice when authorship was hidden [18]. The demand for physician oversight and official certification is the most direct means of steering parents toward safer tools, it held across user categories, and it maps closely onto the digital information platform for parents that the French first-thousand-days commission called for [7].

Two patterns reach beyond conventional information-seeking. For most users, AI was additive rather than substitutive: 66.8% reported no change in how often they consulted a clinician and only 6.9% reported fewer visits, while 18.9% said they came better prepared, a profile that fits earlier findings of parents searching online both before and after consultations [21] and that should reassure clinicians worried about disengagement. The less expected pattern was disclosure: a third of parents had confided in an AI tool something they had withheld from both their clinician and their personal network, most often about their own well-being (25.2%) or doubts about their parenting (10.9%). Disclosure rose sharply with use, from 15% of non-users to 43% of occasional and 64% of regular users, and among regular users it approached half for well-being topics and a quarter for parenting doubts. This pattern is consistent with experimental evidence that people disclose more intimate information to a chatbot than to a person, helped by its anonymity and perceived absence of judgment [25], and with survey evidence that adults in many countries already use generative AI for emotional and mental-health support, including companionship and well-being [26]. For an engaged minority, AI is functioning as a confidant and a pre-consultation outlet rather than only an information source. Whether this relieves unmet emotional need or displaces helpful human contact cannot be settled with these data, and reports that some users feel embarrassed about turning to AI rather than a person suggest the experience is not uniformly positive [26].

### Strengths and Limitations

The principal strengths were the size and timing of the sample and the breadth of what it captured. In all, 12,634 parents took part, more than 5,400 answering the concerns questionnaire and more than 7,200 the literacy-and-AI questionnaire, recruited across France in 2026 through a widely used digital health platform at an early point in the spread of generative AI, when contemporary data on parental use are scarce. The study spanned the period from birth to age four, when health-related decisions and preventive contacts are most frequent, and it reached fathers and families that surveys often miss, including single parents and rural residents.

Several limitations bound the findings. The design was cross-sectional, so no direct causal nor temporal claim could be made, and the reported rise in AI use rests on retrospective self-report, which is open to recall and desirability bias, as are the self-rated confidence and disclosure items. Recruitment through a digital platform favors digitally engaged, urban, and more educated parents, and both samples were overwhelmingly maternal, with mothers making up 94.6% of Questionnaire 1 (294 fathers) and 92.9% of Questionnaire 2 (515 fathers), so comparisons between mothers and fathers are exploratory and may not extend to fathers at large. The concerns-and-confidence questionnaire and the literacy-and-AI questionnaire were answered by different respondents, so associations between informational needs and AI use cannot be examined at the individual level. Within the literacy-and-AI questionnaire, the multivariable model tested whether health literacy predicts AI use, but not whether it shapes how critically parents appraise AI-generated content, which was not measured.

The instruments were built for this study without formal piloting or validation; health literacy was measured with a four-item knowledge quiz rather than a validated scale, and confidence showed a ceiling effect that limits discrimination. Missing data were appreciable on the optional sociodemographic items; descriptive and bivariate analyses used the observed data and the multivariable model used multiple imputation, but the fraction of missing information exceeded 0.4 for two predictors, so those adjusted estimates should be read with caution. Although the overall sample was large and exceeded the target derived from the sample-size calculation, some subgroups remained small, notably fathers (n=294 in Questionnaire 1 and n=515 in Questionnaire 2), so estimates restricted to those subgroups should be read cautiously. The population was French-speaking, resident in France, and recruited from a single platform, which limits generalizability to other settings and to parents who do not use such services.

### Future Directions

Several questions follow directly from these results. Future research should prioritize longitudinal studies to determine if the self-reported rise in AI use is real and sustained, and whether AI keeps complementing professional care or starts to substitute for it. Future investigation should investigate whether higher health literacy enhances the critical appraisal of AI content, rather than merely driving adoption: pairing a validated literacy instrument with an objective test of parents’ ability to detect inaccurate AI advice would address this directly. Additionally, future designs should oversample fathers and underrepresented populations to address the maternal skew observed here. Given that parents currently prioritize physician oversight and official certification, future interventions should evaluate if clinical endorsement and supervised tools can effectively steer parents toward safer resources. The disclosure-to-AI pattern calls for qualitative and mixed-methods analysis of what parents seek when they confide in a conversational agent and of the effects for help-seeking; the free-text responses collected in this study but not analyzed here offer a basis for such work. Finally, future work should explore the dynamics of both health topics interests and AI usage along one full year, all four seasons included, as well as cultural differences across countries.

### Conclusions

This study offers an early, large-scale account of what parents of 0-to-4-year-old children in France want to know, what they know, and how they are beginning to use generative AI for their children’s health in 2026. Unmet informational need is common and concentrated on everyday developmental and behavioral questions, and both confidence and knowledge are shaped less by gender or experience than by education and access to care. Generative AI usage is rising among parents, with over half now incorporating it into their information routine. While most view it as a complement to professional care, an engaged minority treats it as a confidant. Parents consistently condition their trust on physician oversight and official certification. Because higher health literacy was associated with greater AI use, there is a risk that these tools will widen the information gap rather than close it, since the parents who are already best informed adopt it most. These findings highlight the need for clinically anchored, certified digital resources designed with the social gradient in mind, ensuring that AI adoption helps reduce, rather than widen, the inequalities that take root in a child’s first years.

## Data Availability

The individual-level data are not publicly available. Although the questionnaire collected no direct identifiers and recorded sociodemographic variables only in broad categories, participants were not informed that their responses would be deposited publicly, and open release would therefore exceed the commitments made to them. Aggregated data supporting the findings are presented within the manuscript and its multimedia appendices. The analysis code is publicly available at https://doi.org/10.5281/zenodo.22703273.

## Acknowledgments

The authors thank the parents who took part in the study for their time and their trust. They also thank the Doctolib teams who supported the project, in particular the communications team, who helped design and disseminate the study invitation, and the legal and data-protection team, who advised on the regulatory and privacy aspects of the survey.

During the preparation of this manuscript, the authors used Claude (Anthropic) to assist with language editing and with formatting the manuscript, with the statistical analysis, and with the interpretation of the results. Generative AI was not used for the study design and data collection. The authors reviewed, verified, and edited all AI-assisted content and take full responsibility for the integrity and accuracy of the manuscript.

## Data Availability

The individual-level data generated and analyzed in this study are not publicly available. They consist of survey responses containing sociodemographic information, collected under the data-protection commitments made to participants, and sharing them would be incompatible with those commitments. Aggregated data supporting the findings are presented within the manuscript and its multimedia appendices. The analysis code is publicly available at https://doi.org/10.5281/zenodo.22703273

## Authors’ Contributions

KG: Conceptualization, Data curation, Investigation, Methodology, Project administration, Supervision, Validation, Visualization, Writing - original draft, Writing - review & editing.

NB: Data curation, Formal analysis, Software, Validation, Visualization, Writing - review & editing. A-KEB: Conceptualization, Formal analysis, Supervision, Writing - review & editing.

PM: Conceptualization, Methodology, Project administration, Supervision, Writing - original draft, Writing - review & editing.

All authors made substantial contributions, critically reviewed the manuscript, approved the final version, and agree to be accountable for all aspects of the work, in accordance with the ICMJE criteria.

## Abbreviations

AI: artificial intelligence
CI: confidence interval
GEE: generalized estimating equations
HCP: health care professional
IQR: interquartile range
OR: odds ratio
SD: standard deviation

## Multimedia Appendices

**Multimedia Appendix 1.**
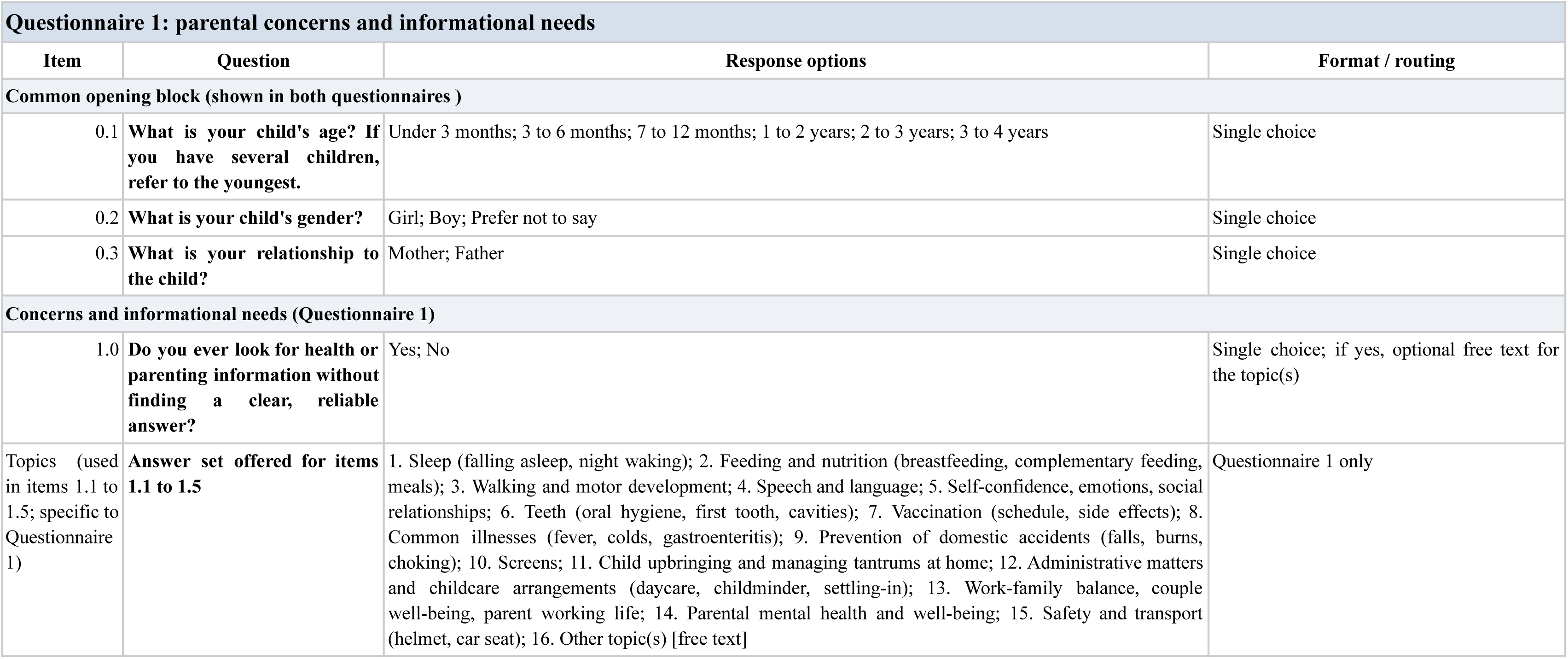
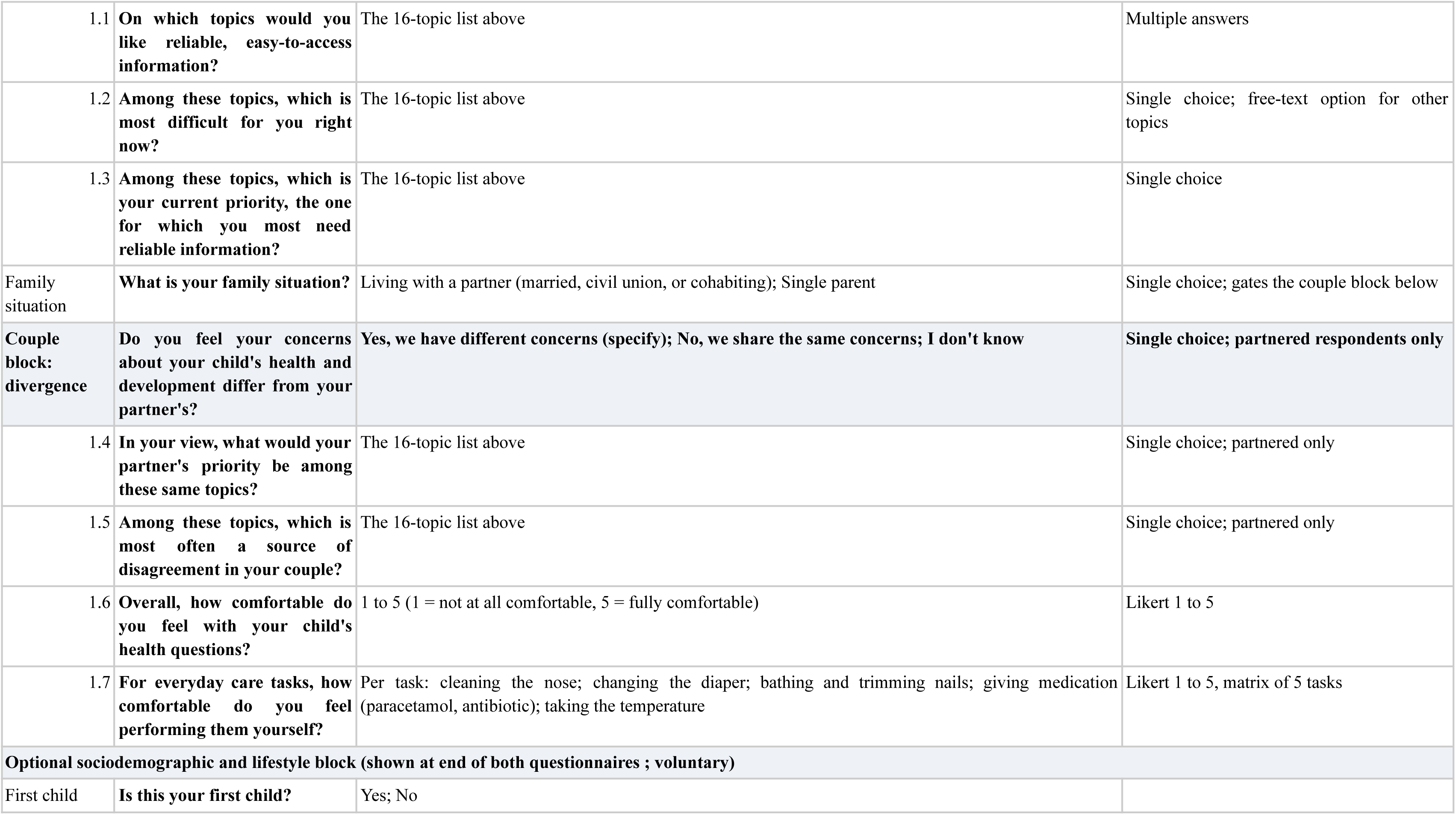
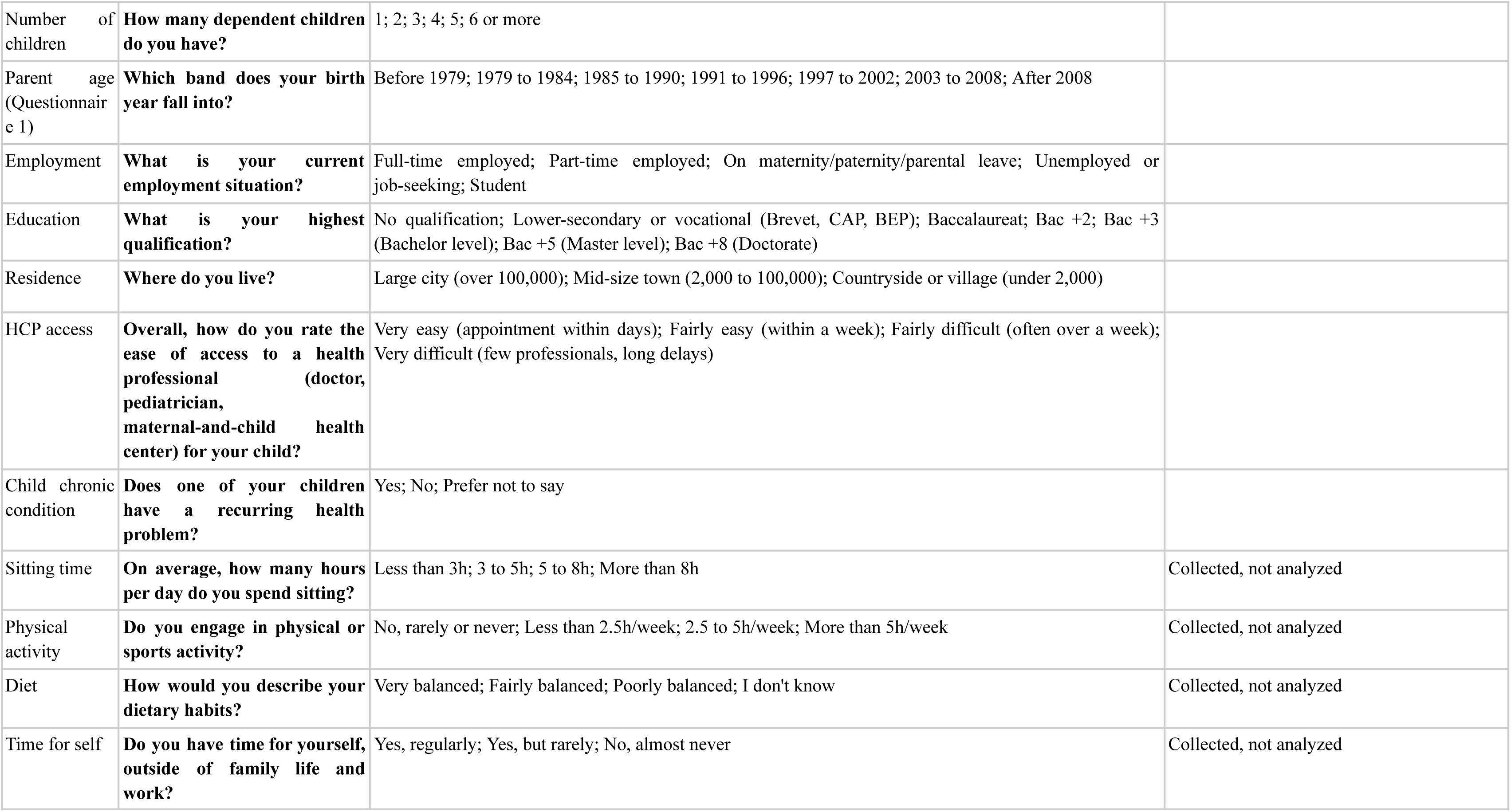
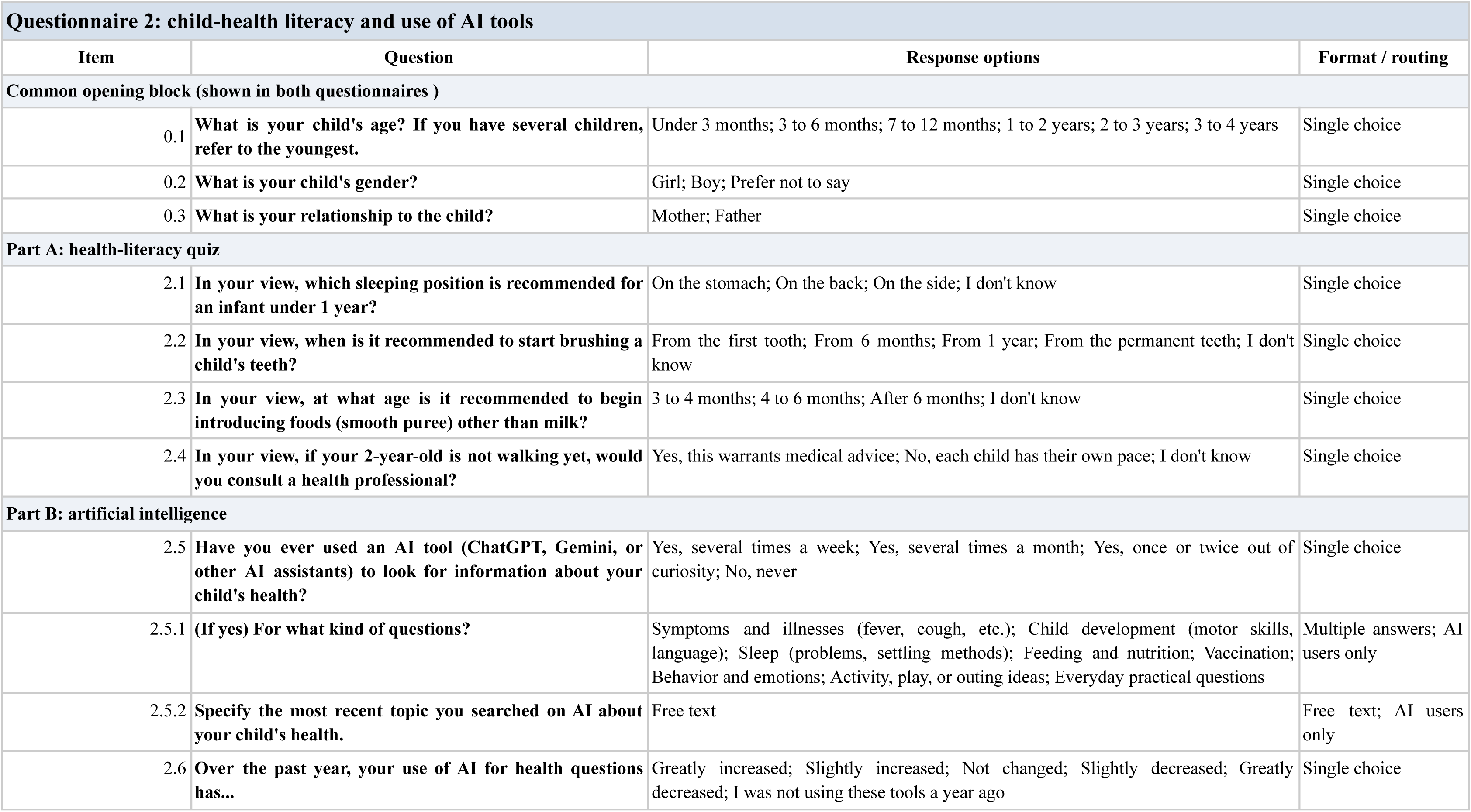
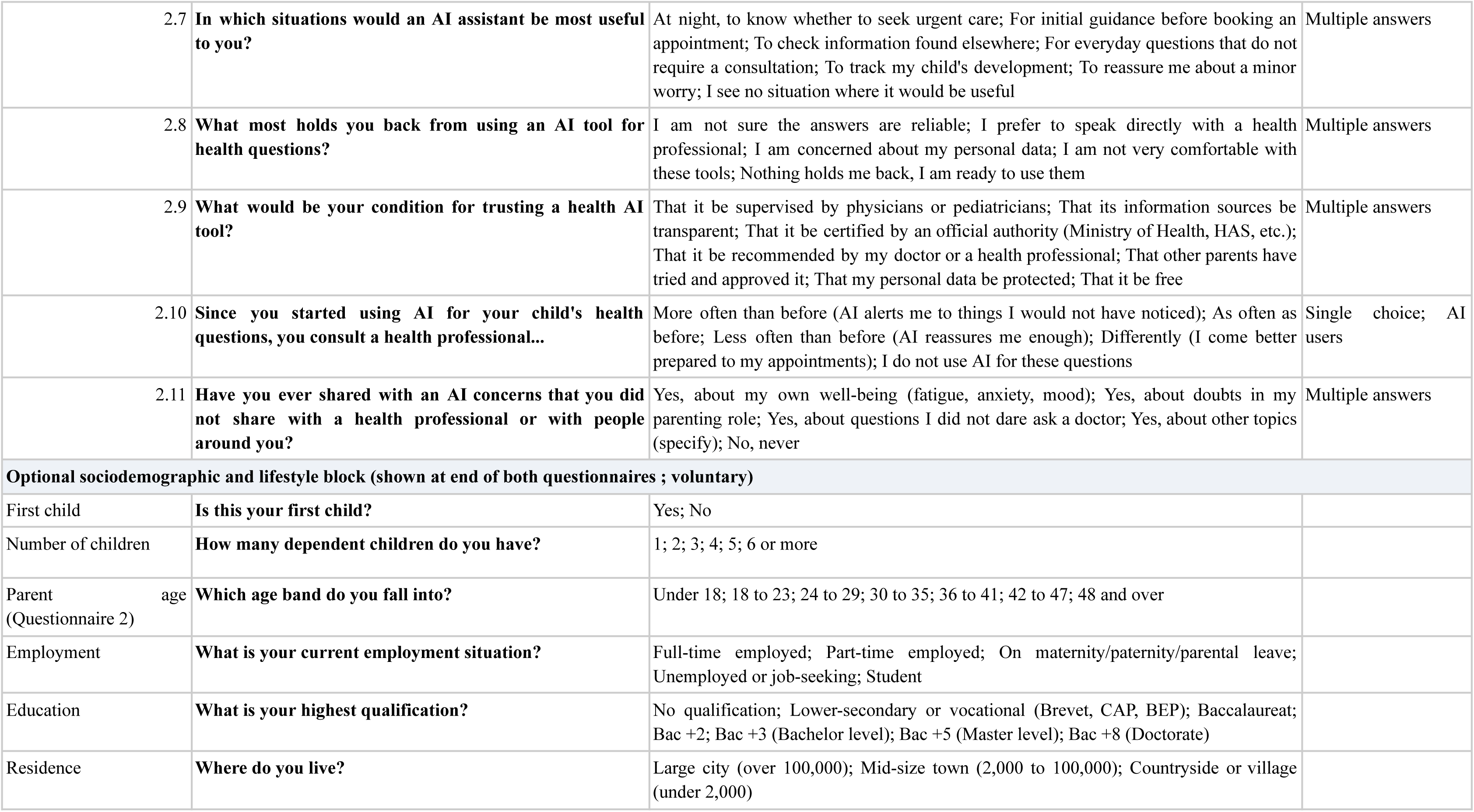
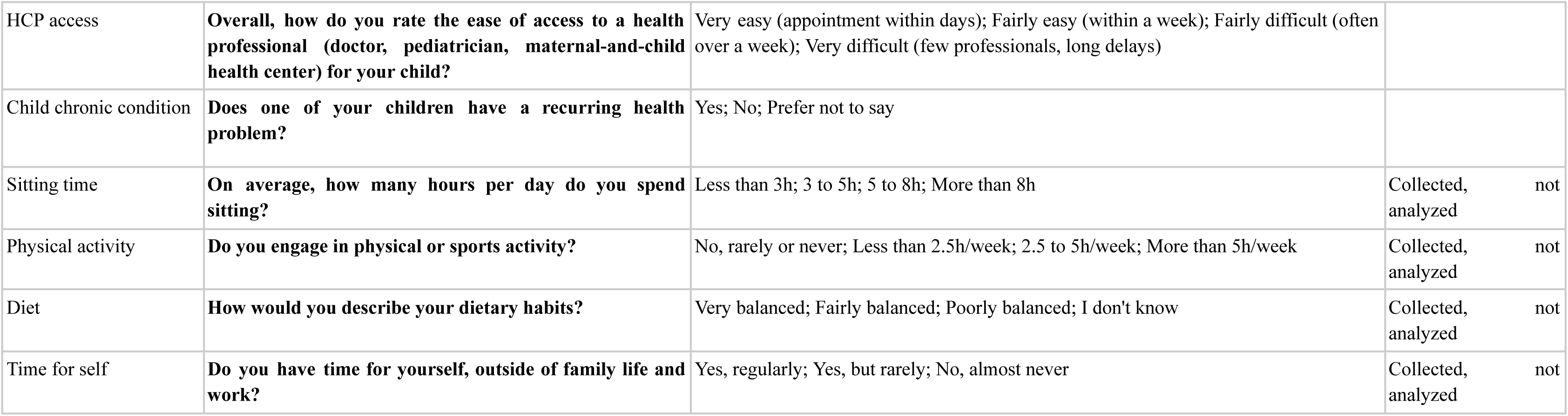
Study instrument (Questionnaire 1 and Questionnaire 2, English translation). Full text of the two self-administered questionnaires, with item wording, response options, and display conditions. Both questionnaires open with three items on the child and respondent and close with an optional sociodemographic and lifestyle block. Questionnaire 1 covers informational needs and concerns, confidence in daily care, and, for partnered respondents, perceived agreement with the partner; Questionnaire 2 covers child-health knowledge and the use and perception of AI tools. Translated from the original French.

**Multimedia Appendix 2.**
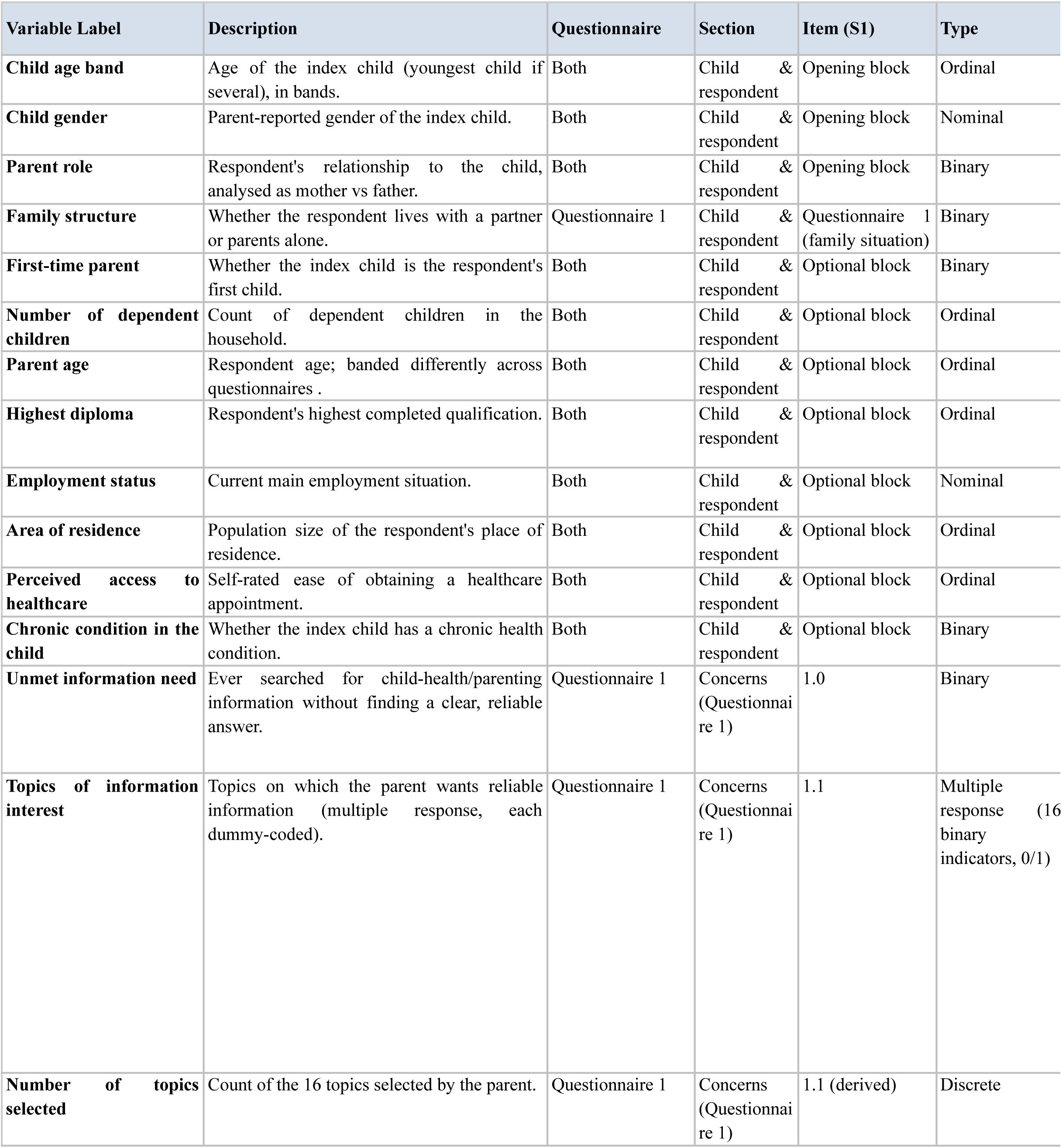
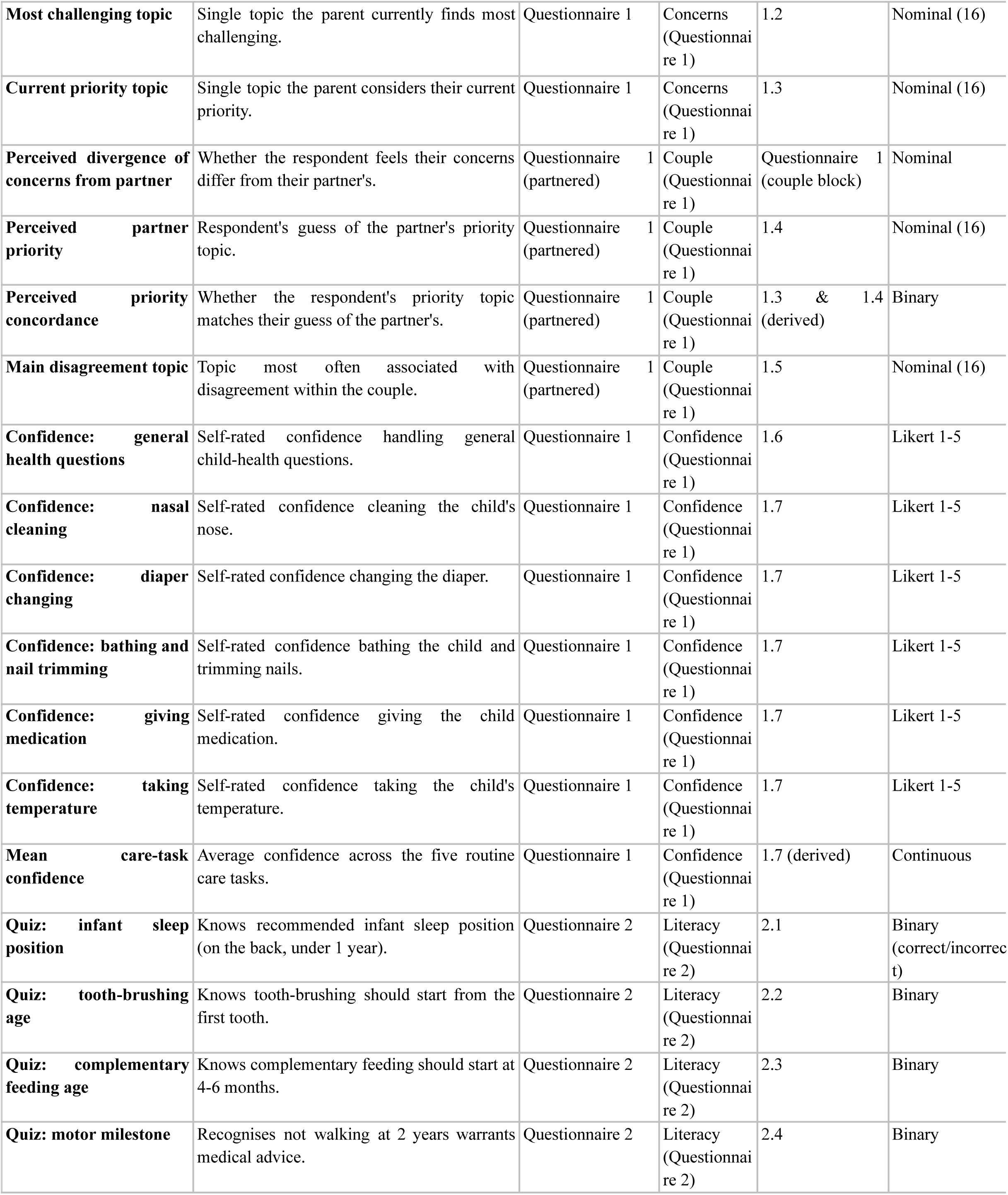
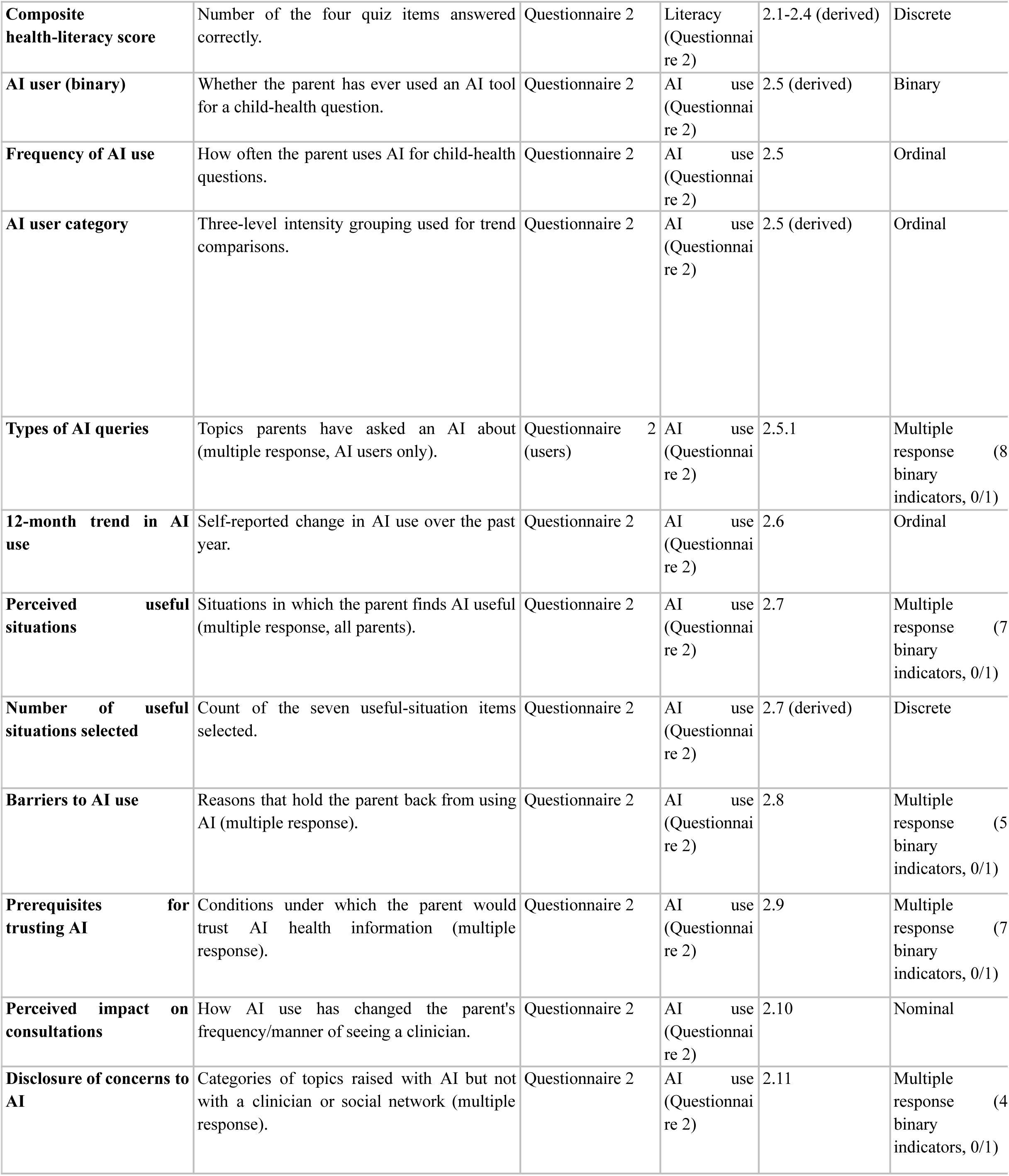
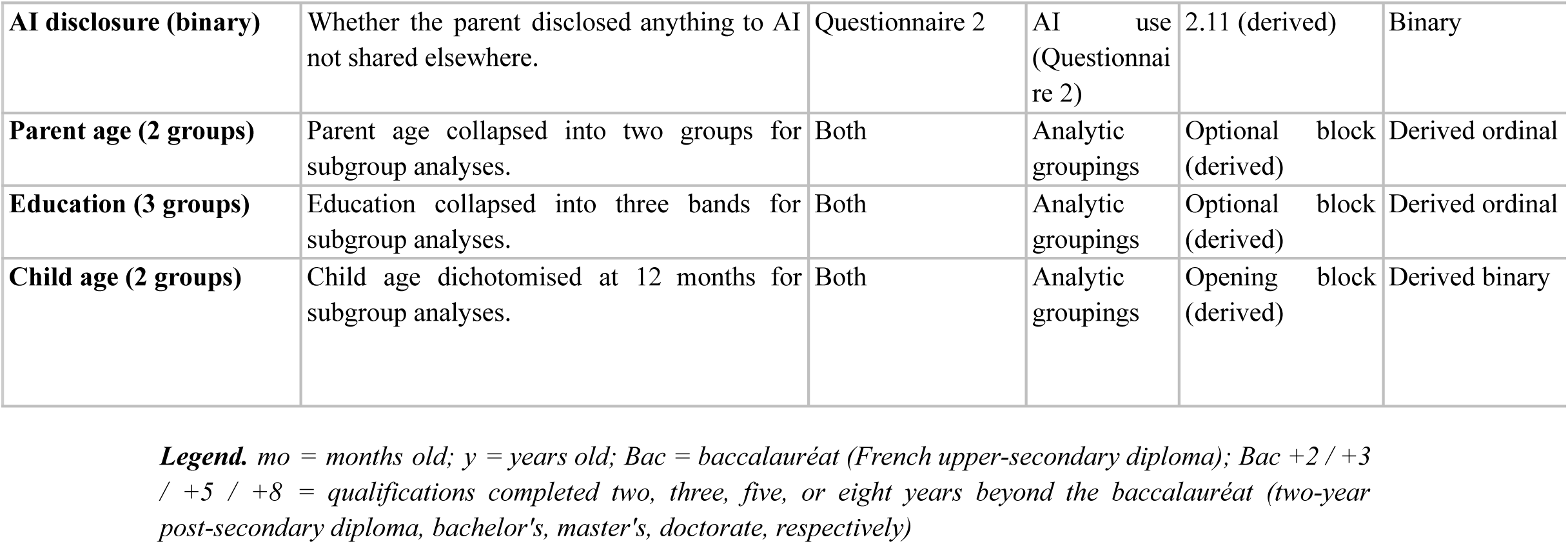
Data dictionary. Every analyzed variable with its label, description, source questionnaire and section, source questionnaire item, type, response options, derivation, and the table or figure in which it appears.

**Multimedia Appendix 3.** Checklist for Reporting Results of Internet E-Surveys (CHERRIES)

| Checklist item | Response (INFANT study) |
| --- | --- |
| **Design** |  |
| **Survey design** | Cross-sectional online survey using a convenience sample. Two self-administered questionnaires (Questionnaire 1 on parental concerns and information needs; Questionnaire 2 on child-health literacy and use of AI) were disseminated through a marketing card in the Doctolib application between May 12 and June 11, 2026, to French-speaking parents of a child aged 0 to 47 months residing in France. Open survey. |
| **IRB approval and informed consent process** |  |
| **IRB approval** | No approval by a Committee for the Protection of Persons (Comité de Protection des Personnes) was required: as an observational, non-interventional survey, the study fell outside the scope of the French Jardé Law (see Ethics and Data Protection) and outside of the General Data Protection Regulation (GDPR). |
| **Informed consent** | No informed consent is required for this study as it falls outside the scope of the GDPR, the French Data Protection Act (Loi Informatique et Libertés) and the French framework for research involving the human person (Loi n° 2012-300 du 5 mars 2012 relative aux recherches impliquant la personne humaine, known as the Loi Jardé).<br>Before starting, participants were informed through a short notice describing the purpose of the questionnaire, stating that responses were anonymous, and linking to Doctolib's personal-data protection policy regarding the possibility to push forth voluntary questionnaires. Participation was entirely voluntary. No personal data was collected as part of the questionnaire. |
| **Data protection** | The study falls outside of the scope of the GDPR and the French Data Protection Act. No personal data was collected. Before starting, participants were shown a short notice describing the purpose of the questionnaire, stating that responses were anonymous, and linking to Doctolib's personal-data protection policy regarding the possibility to push forth voluntary questionnaires under Doctolib's legitimate interest. The questionnaire was designed to collect only anonymous, non-identifying information, such as general responses, age bands, and broad categories for other variables. No directly identifying information was requested. Each free-text field included a reminder asking participants not to enter information that could identify them. The responses themselves did not constitute personal data within the meaning of the GDPR. Responses were exported from Typeform to Doctolib servers certified for health-data hosting (Hébergeur de Données de Santé, HDS), with access restricted to the research team; all analyses were performed on aggregated data. |
| **Development and pre-testing** |  |
| **Development and testing** | The questionnaires were developed from a review of the literature on parental health literacy, early-childhood health, parental information needs, and the use of AI tools in health care. No formal pilot testing or usability/technical testing was carried out before fielding. |
| **Recruitment process and description of the sample having access to the questionnaire** |  |
| **Open survey versus closed survey** | Open survey. Accessible to users of the Doctolib application in the target group, through a marketing card. |
| **Contact mode** | Contact was made online. Participants were not individually solicited; they were exposed to a marketing card within the Doctolib application. |
| **Advertising the survey** | The survey was advertised through a marketing card within the Doctolib application; no other channel was used. The full questionnaire wording is provided in Multimedia Appendix 1. |
| **Survey administration** |  |
| **Web/E-mail** | Web-based survey hosted on Typeform and accessed from within the Doctolib application; responses were captured automatically. |
| **Context** | The survey was displayed within the Doctolib application, a widely used online health-care platform in France whose users manage health-care appointments and related tasks. The health-care context of the platform may have pre-selected respondents already engaged with digital health. |
| **Mandatory/voluntary** | Voluntary. |
| **Incentives** | No incentive or reward was offered. |
| **Time/Date** | May 12 to June 11, 2026. |
| **Randomization of items or questionnaires** | Items were presented in a fixed order; no randomization was used. |
| **Adaptive questioning** | Yes. Some items were displayed conditionally (eg, the couple block was shown only to respondents living with a partner; AI-use details were shown only to respondents who reported using AI). |
| **Number of items** | The questionnaires used branching and matrix items, so a single item count would be misleading; the instrument is described by domain. The full set of items is provided in Multimedia Appendix 1. |
| **Number of screens (pages)** | One item per screen (Typeform); the effective number of screens varied with branching. |
| **Completeness check** | Yes. Mandatory items (marked with an asterisk) had to be answered before moving to the next item; other items were optional. |
| **Review step** | Yes. Respondents could go back and change their answers before submitting. |
| **Response rates** |  |
| **Unique site visitor** | Uniqueness was based on the survey platform's visitor counting. First-page views (survey visitors): 24,422 for Questionnaire 1 and 23,622 for Questionnaire 2. The number of users to whom the marketing card was shown (unique site visitors/impressions) was not available. |
| **View rate** | Not available. Computing a view rate would require the number of card impressions (unique site visitors), a platform metric that was not available. |
| **Participation rate** | Starts divided by first-page views. Questionnaire 1: $10,987/24,422 = 45.0\%$. Questionnaire 2: $11,275/23,622 = 47.7\%$. |
| **Completion rate** | Valid submitted questionnaires divided by starts. Questionnaire 1: $5,410/10,987 = 49.2\%$. Questionnaire 2: $7,224/11,275 = 64.1\%$. |
| **Preventing multiple entries from the same individual** |  |
| **Cookies used** | No. |
| **IP check** | No. |
| **Log file analysis** | No. |
| **Registration** | The survey was accessed from within the Doctolib application by logged-in users, but no restriction to a single response per account was enforced, and no registration-based deduplication was applied. |
| **Analysis** |  |
| **Handling of incomplete questionnaires** | Only fully submitted questionnaires were received: submission was possible only after all mandatory items had been answered, so partially completed questionnaires were not captured. The analyzed samples were 5,410 (Questionnaire 1) and 7,224 (Questionnaire 2). Item-level missingness, confined to the optional blocks, was handled by multiple imputation by chained equations (20 imputations, Rubin's rules) for the multivariable model. |
| **Questionnaires submitted with an atypical timestamp** | Completion times were not analyzed, and no responses were excluded on this basis. |
| **Statistical correction** | No weighting or post-stratification was applied; the non-representativeness of the sample is addressed as a limitation in the Discussion. |

---

**Multimedia Appendix 4.**
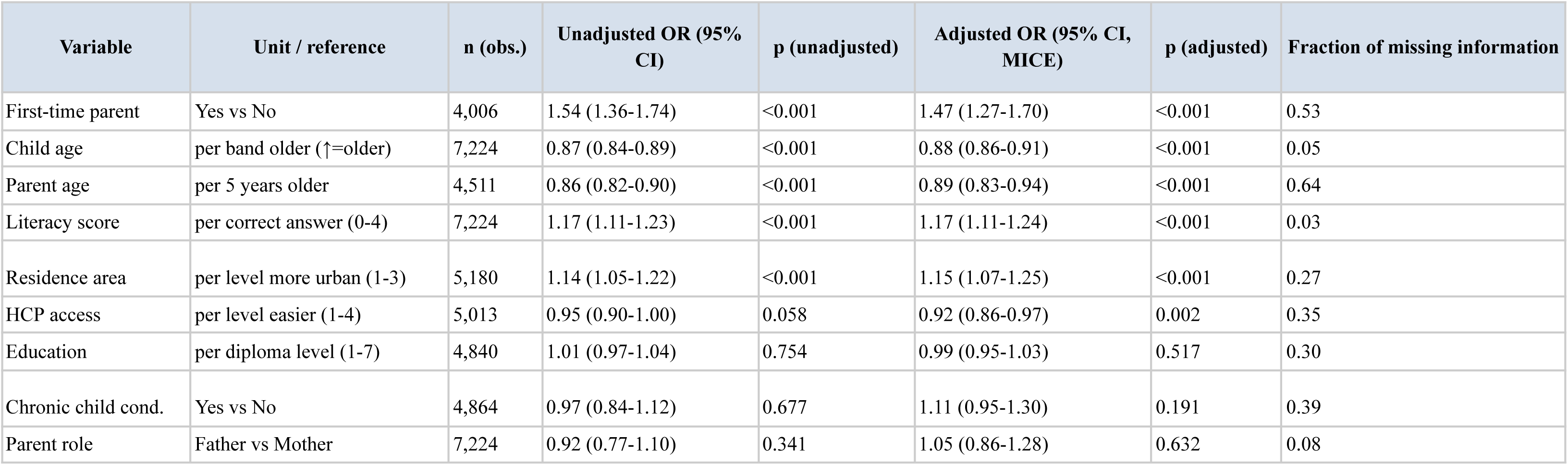
Predictors of AI use: univariate and multivariable logistic regression. Questionnaire 2 (n = 7,224). Unadjusted ORs from univariate logistic regression (complete cases per variable). Adjusted ORs from multivariable logistic regression pooled across 20 MICE-imputed datasets (Rubin’s rules). FMI = fraction of missing information; values above 0.40 (first-time parenthood, parent age) indicate greater sensitivity to the imputation model. Employment status omitted from the regression model (nominal 5-level categorical, 29% missing; see Multimedia *Appendix 5*). Numerical values underlying *Figure 2*.

**Multimedia Appendix 5.**
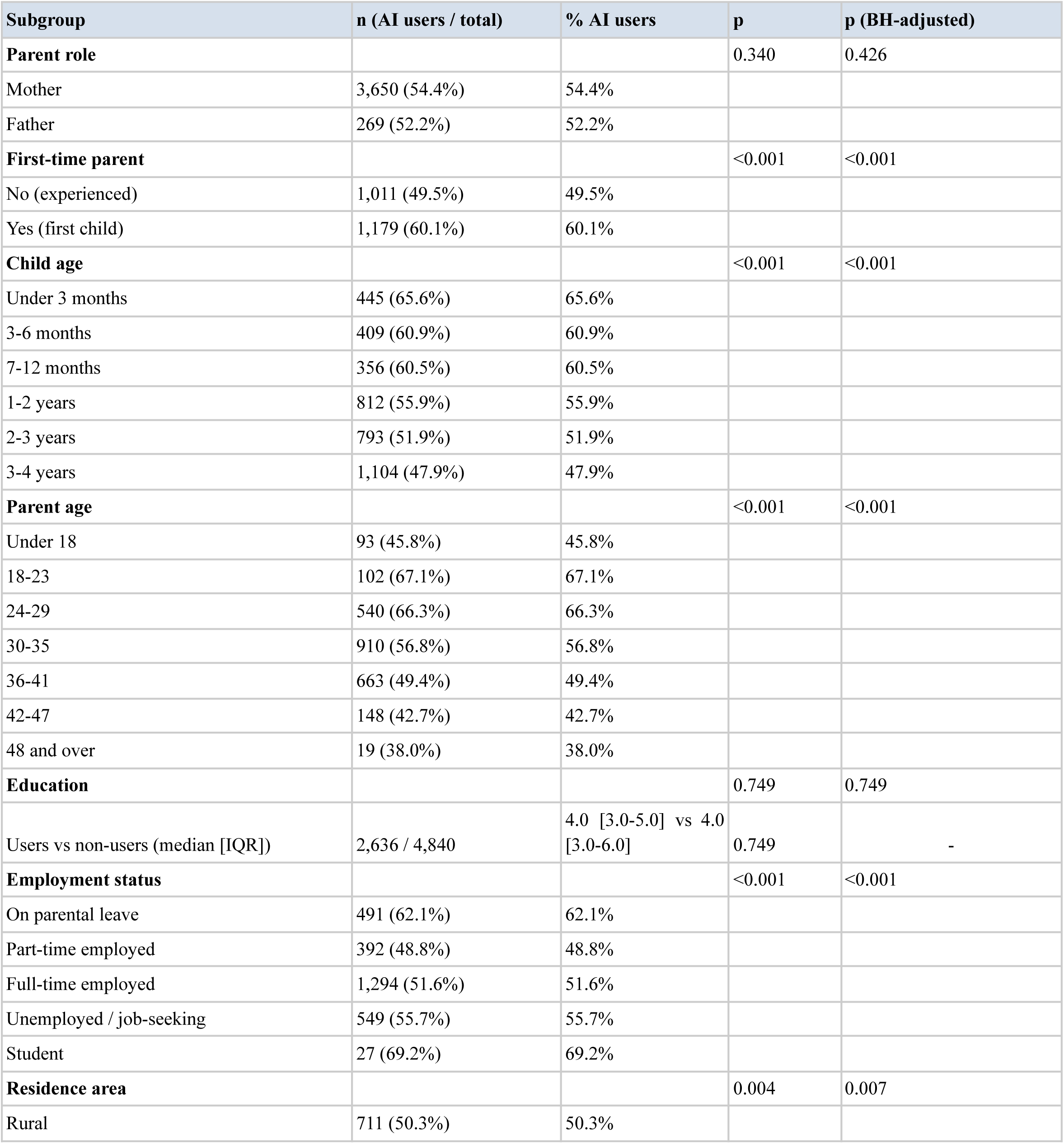
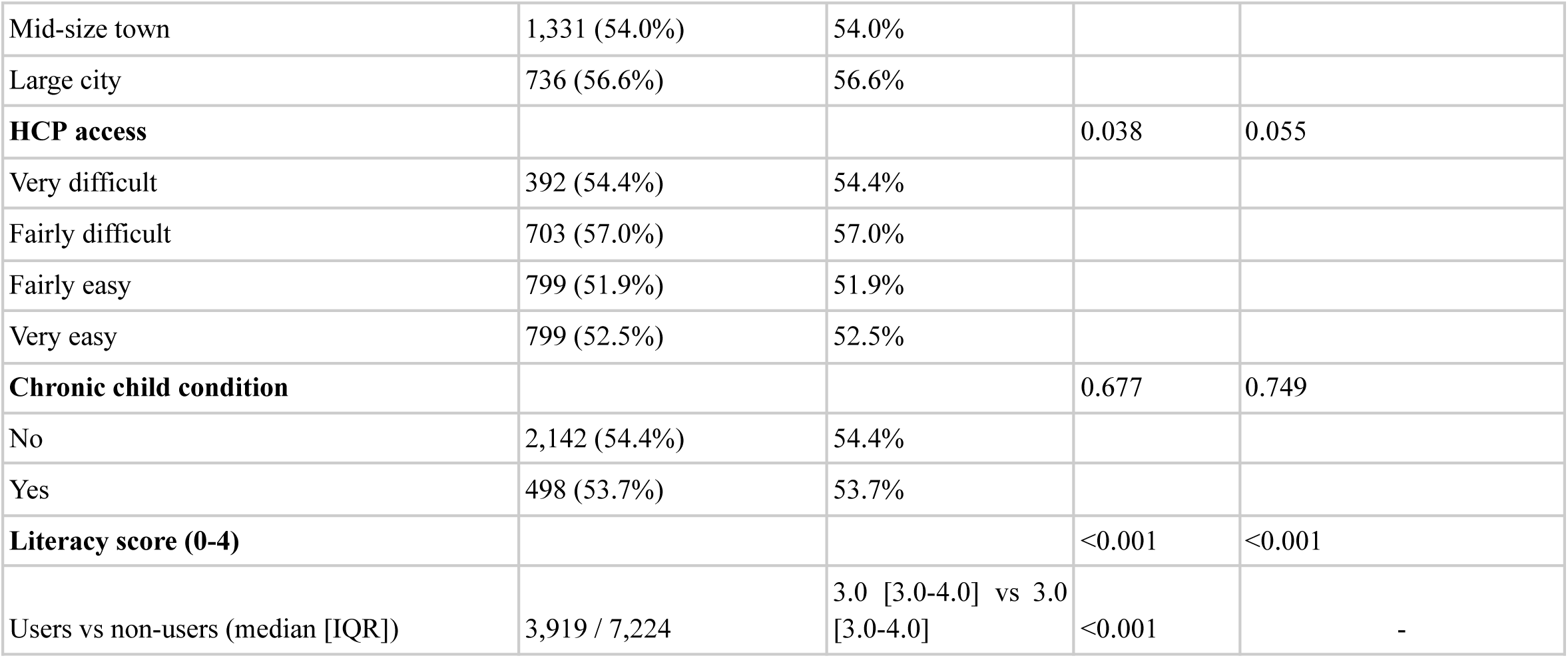
Proportion of AI users by sociodemographic subgroup, unadjusted. Questionnaire 2 (n = 7,224). Unadjusted proportion of AI users within each subgroup level. Categorical variables by chi-square with Fisher fallback; ordinal variables (education and literacy score) by Mann-Whitney U. Raw and Benjamini-Hochberg-adjusted p-values (p adj.) across ten tests are reported. Adjusted estimates appear in *Figure 2*.

**Multimedia Appendix 6.** Informational needs and concerns (full version of Table 2)

| Topic | Overall (selected) | Most challenging | Priority | Mother | Father | p (M vs F) | p adj. (M vs F) | First-time | Experienced | p (parity) | p adj. (parity) |
| --- | --- | --- | --- | --- | --- | --- | --- | --- | --- | --- | --- |
| Sleep | 2,776 (51.3%) | 1,613 (29.8%) | 1,390 (25.7%) | 2,631 (51.4%) | 145 (49.3%) | 0.520 | 0.583 | 1,238 (57.2%) | 1,013 (47.6%) | <0.001 | <0.001 |
| Nutrition | 2,600 (48.1%) | 758 (14.0%) | 848 (15.7%) | 2,473 (48.3%) | 127 (43.2%) | 0.098 | 0.261 | 1,253 (57.9%) | 896 (42.1%) | <0.001 | <0.001 |
| Education | 2,587 (47.8%) | 688 (12.7%) | 746 (13.8%) | 2,481 (48.5%) | 106 (36.1%) | **<0.001** | <0.001 | 1,082 (50.0%) | 1,060 (49.8%) | 0.926 | 0.976 |
| Self-confidence & emotions | 2,429 (44.9%) | 466 (8.6%) | 436 (8.1%) | 2,321 (45.4%) | 108 (36.7%) | **0.005** | 0.015 | 1,004 (46.4%) | 962 (45.2%) | 0.453 | 0.604 |
| Common illnesses | 2,138 (39.5%) | 246 (4.5%) | 257 (4.8%) | 2,049 (40.1%) | 89 (30.3%) | **0.001** | 0.004 | 1,004 (46.4%) | 746 (35.1%) | <0.001 | <0.001 |
| Parental mental health | 2,064 (38.2%) | 174 (3.2%) | 188 (3.5%) | 1,980 (38.7%) | 84 (28.6%) | **<0.001** | 0.003 | 862 (39.8%) | 854 (40.1%) | 0.867 | 0.976 |
| Language | 1,951 (36.1%) | 380 (7.0%) | 472 (8.7%) | 1,852 (36.2%) | 99 (33.7%) | 0.415 | 0.583 | 823 (38.0%) | 763 (35.9%) | 0.148 | 0.216 |
| Dental hygiene | 1,898 (35.1%) | 210 (3.9%) | 182 (3.4%) | 1,803 (35.2%) | 95 (32.3%) | 0.337 | 0.583 | 947 (43.8%) | 587 (27.6%) | <0.001 | <0.001 |
| Work-family balance | 1,790 (33.1%) | 261 (4.8%) | 253 (4.7%) | 1,701 (33.2%) | 89 (30.3%) | 0.322 | 0.583 | 786 (36.3%) | 690 (32.4%) | 0.008 | 0.013 |
| Accident prevention | 1,404 (26.0%) | 64 (1.2%) | 83 (1.5%) | 1,356 (26.5%) | 48 (16.3%) | **<0.001** | 0.001 | 654 (30.2%) | 505 (23.7%) | <0.001 | <0.001 |
| Screens | 1,351 (25.0%) | 145 (2.7%) | 153 (2.8%) | 1,273 (24.9%) | 78 (26.5%) | 0.572 | 0.583 | 609 (28.1%) | 485 (22.8%) | <0.001 | <0.001 |
| Vaccination | 1,278 (23.6%) | 62 (1.1%) | 62 (1.1%) | 1,220 (23.8%) | 58 (19.7%) | 0.122 | 0.279 | 508 (23.5%) | 513 (24.1%) | 0.652 | 0.803 |
| Motor development | 1,038 (19.2%) | 114 (2.1%) | 152 (2.8%) | 988 (19.3%) | 50 (17.0%) | 0.368 | 0.583 | 533 (24.6%) | 324 (15.2%) | <0.001 | <0.001 |
| Childcare | 796 (14.7%) | 65 (1.2%) | 67 (1.2%) | 749 (14.6%) | 47 (16.0%) | 0.583 | 0.583 | 397 (18.3%) | 259 (12.2%) | <0.001 | <0.001 |
| Safety & transport | 678 (12.5%) | 25 (0.5%) | 18 (0.3%) | 645 (12.6%) | 33 (11.2%) | 0.545 | 0.583 | 327 (15.1%) | 234 (11.0%) | <0.001 | <0.001 |
| Other | 254 (4.7%) | 138 (2.6%) | 102 (1.9%) | 237 (4.6%) | 17 (5.8%) | 0.445 | 0.583 | 97 (4.5%) | 94 (4.4%) | 0.976 | 0.976 |

**Multimedia Appendix 7.** Per-topic odds ratios for topic selection, mother versus father. Questionnaire 1 (n = 5,410; mothers = 5,116, fathers = 294). OR = odds ratio (mother vs father; father = reference); OR > 1 means mothers were more likely to flag the topic. 95% CIs from non-parametric bootstrap (2,000 resamples, percentile method). p: chi-square (Fisher for small 2×2). p (BH-adjusted): Benjamini-Hochberg FDR across the 16 tests. GEE topic-by-parent-role interaction (binomial, exchangeable working correlation, clustered by respondent): p < 0.001. Numerical values underlying Multimedia *Appendix 8*.

| Topic | OR (mother vs father) | 95% CI | p (raw) | p (BH-adjusted) |
| --- | --- | --- | --- | --- |
| Accident prevention | 1.85 | 1.38-2.59 | <0.001 | 0.001 |
| Education | 1.67 | 1.31-2.12 | <0.001 | <0.001 |
| Parental mental health | 1.58 | 1.24-2.06 | <0.001 | 0.003 |
| Common illnesses | 1.54 | 1.21-1.98 | 0.001 | 0.004 |
| Self-confidence & emotions | 1.43 | 1.11-1.81 | 0.005 | 0.015 |
| Vaccination | 1.27 | 0.96-1.73 | 0.122 | 0.279 |
| Other | 0.79 | 0.52-1.52 | 0.445 | 0.583 |
| Nutrition | 1.23 | 0.98-1.56 | 0.098 | 0.261 |
| Motor development | 1.17 | 0.87-1.62 | 0.368 | 0.583 |
| Work-family balance | 1.15 | 0.89-1.48 | 0.322 | 0.583 |
| Safety & transport | 1.14 | 0.80-1.70 | 0.545 | 0.583 |
| Dental hygiene | 1.14 | 0.88-1.48 | 0.337 | 0.583 |
| Language | 1.12 | 0.87-1.45 | 0.415 | 0.583 |
| Childcare | 0.90 | 0.66-1.28 | 0.583 | 0.583 |
| Screens | 0.92 | 0.70-1.21 | 0.572 | 0.583 |
| Sleep | 1.09 | 0.86-1.38 | 0.520 | 0.583 |

**Multimedia Appendix 8.**
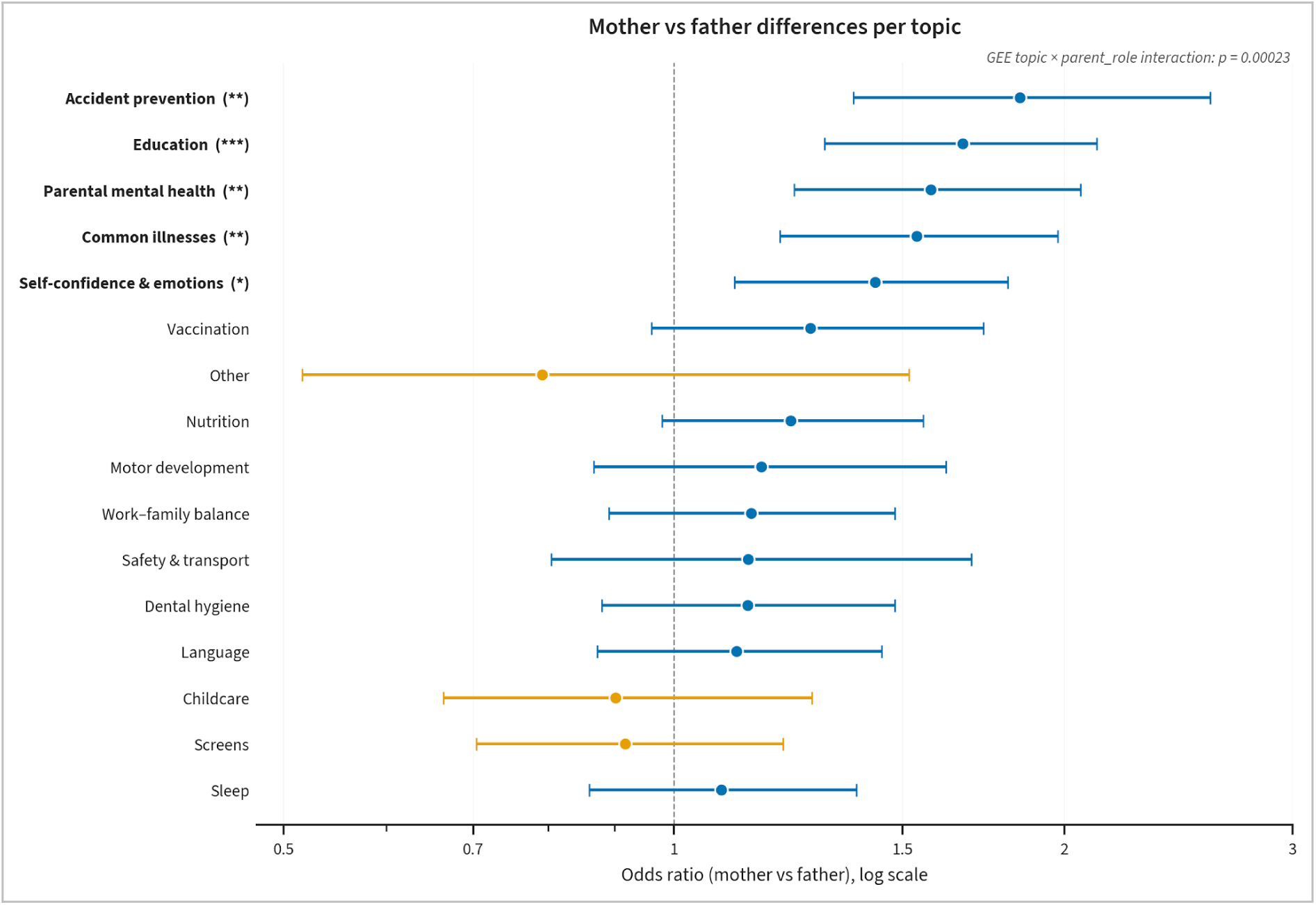
Per-topic odds ratios (mother vs father), forest plot. Questionnaire 1 (n=5,410; mothers=5,116, fathers=294). Odds ratios from per-topic univariate logistic regression, with 95% confidence intervals from nonparametric bootstrap (2,000 resamples). The x-axis is on a logarithmic scale; an odds ratio above 1 indicates a topic selected more often by mothers, below 1 more often by fathers. The generalized estimating equations omnibus test for the topic-by-parental-role interaction is annotated. Numerical values are given in Multimedia *Appendix 7*. Significance (Benjamini-Hochberg-adjusted): *P<.05, **P<.01, ***P<.001.

**Multimedia Appendix 9.**
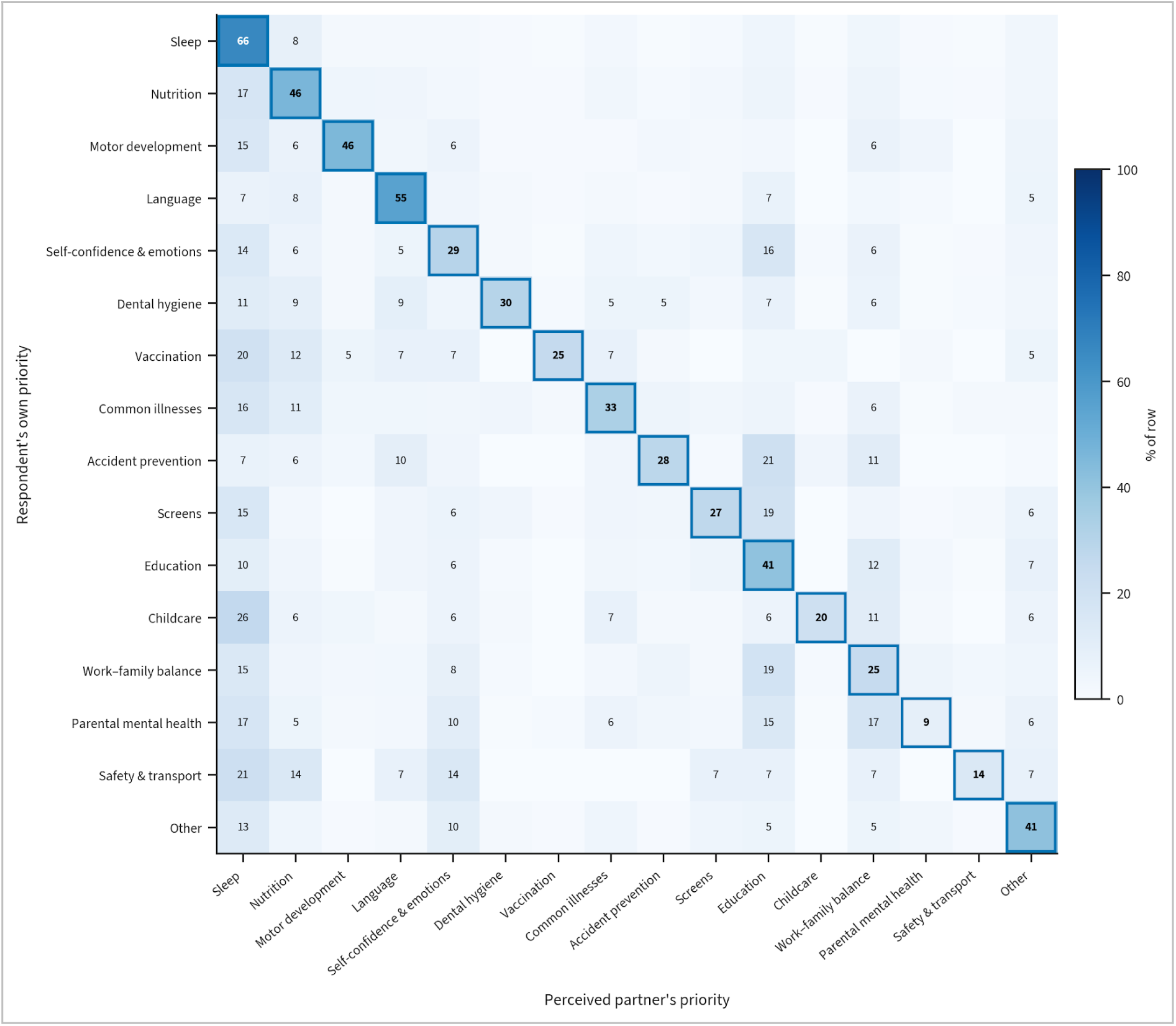
Perceived intra-couple priority concordance, heatmap. Questionnaire 1 (coupled respondents n=4,544; respondents with both items answered n=4,543). Row-normalized heatmap: rows are the respondent’s own priority topic, columns the priority attributed to the partner; diagonal cells indicate perceived agreement (Cohen κ=0.368). No paired couple data were collected, so the figure reflects the respondent’s perception of the partner’s priority, not the partner’s own answer.

**Multimedia Appendix 10.** Confidence in general health questions and daily care tasks. Questionnaire 1 (n = 5,410; mothers = 5,116, fathers = 294). Confidence rated from 1 (not at all confident) to 5 (fully confident).The composite confidence score is the mean of the five care-task items (cleaning the child’s nose, changing the diaper, bathing and trimming nails, giving medication, and taking the temperature); general health confidence is reported separately and is not included in the composite. Mother versus father and first-time versus experienced comparisons by Mann-Whitney U. Significance (Benjamini-Hochberg-adjusted): *P<.05, **P<.01, ***P<.001

| Item | n | Mean (SD) | Median [IQR] | Mother, mean (SD) | Father, mean (SD) | p (M vs F) | First-time (mean ± SD) | Experienced (mean ± SD) | p (parity) | p adj. (M vs F) | p adj. (parity) |
| --- | --- | --- | --- | --- | --- | --- | --- | --- | --- | --- | --- |
| General health confidence | 5410 | 4.12 (0.84) | 4.0 [4.0-5.0] | 4.12 (0.84) | 4.18 (0.81) | 0.237 | 4.03 (0.85) | 4.22 (0.79) | <0.001 | 0.331 | <0.001 |
| Cleaning the child's nose | 5410 | 4.26 (1.05) | 5.0 [4.0-5.0] | 4.26 (1.05) | 4.33 (0.99) | 0.328 | 4.14 (1.13) | 4.37 (0.97) | <0.001 | 0.383 | <0.001 |
| Changing the diaper | 5410 | 4.92 (0.39) | 5.0 [5.0-5.0] | 4.92 (0.37) | 4.79 (0.58) | <0.001 | 4.92 (0.37) | 4.93 (0.37) | 0.265 | <0.001 | 0.265 |
| Bathing and trimming nails | 5410 | 4.68 (0.69) | 5.0 [5.0-5.0] | 4.69 (0.67) | 4.45 (0.88) | <0.001 | 4.62 (0.72) | 4.74 (0.64) | <0.001 | <0.001 | <0.001 |
| Giving medication | 5147 | 4.52 (0.86) | 5.0 [4.0-5.0] | 4.52 (0.86) | 4.54 (0.84) | 0.611 | 4.40 (0.93) | 4.63 (0.75) | <0.001 | 0.611 | <0.001 |
| Taking the temperature | 5320 | 4.72 (0.66) | 5.0 [5.0-5.0] | 4.72 (0.66) | 4.67 (0.70) | 0.135 | 4.66 (0.71) | 4.77 (0.61) | <0.001 | 0.236 | <0.001 |
| Composite confidence | 5410 | 4.62 (0.51) | 4.8 [4.4-5.0] | 4.62 (0.51) | 4.55 (0.58) | 0.081 | 4.55 (0.54) | 4.69 (0.47) | <0.001 | 0.190 | <0.001 |

**Multimedia Appendix 11.** Confidence in daily care tasks, by sociodemographic subgroup. Questionnaire 1 (n = 5,410). Subgroup comparisons by Kruskal-Wallis (ordinal variables with three or more levels) and Mann-Whitney U (binary variables). Cells show BH-FDR-adjusted p adj.-values within the family of seven items. Significance: * p adj. < 0.05, ** p adj. < 0.01, *** p adj. < 0.001.

| Item | Education level | Residence | HCP access | Parent age | Chronic child |
| --- | --- | --- | --- | --- | --- |
| General health confidence | <0.001 *** | 0.269 | <0.001 *** | 0.023 * | 0.191 |
| Cleaning the child's nose | <0.001 *** | 0.393 | <0.001 *** | 0.910 | 0.042 * |
| Changing the diaper | 0.433 | 0.638 | 0.014 * | 0.524 | 0.638 |
| Bathing and trimming nails | <0.001 *** | 0.018 * | <0.001 *** | 0.515 | 0.019 * |
| Giving medication | <0.001 *** | 0.018 * | <0.001 *** | 0.391 | <0.001 *** |
| Taking the temperature | <0.001 *** | 0.010 ** | <0.001 *** | 0.785 | 0.019 * |
| Composite confidence | <0.001 *** | 0.010 ** | <0.001 *** | 0.600 | 0.001 ** |

**Multimedia Appendix 12.** Child-health literacy quiz, by parent role and parity. Questionnaire 2 (n = 7,224). Four binary knowledge items and the composite score (0 to 4 correct answers; mean 3.19, SD 0.91; median 3 [IQR 3-4]). Per-item comparisons by chi-square with Fisher fallback; composite by Mann-Whitney U. Raw and Benjamini-Hochberg adjusted p-values (p adj.) are reported for the mother-versus-father and parity comparisons. Significance (Benjamini-Hochberg-adjusted): *P<.05, **P<.01, ***P<.001

| Item | n | % correct | Mother | Father | p (M vs F) | First-time | Experienced | p (parity) | p (M vs F, adj.) | p (parity, adj.) |
| --- | --- | --- | --- | --- | --- | --- | --- | --- | --- | --- |
| Sleep position (under 1 yr) | 7224 | 6,449 (89.3%) | 6,056 (90.3%) | 393 (76.3%) | <0.001 | 1,795 (91.5%) | 1,824 (89.2%) | 0.018 | <0.001 | 0.074 |
| First tooth-brushing age | 7224 | 4,942 (68.4%) | 4,683 (69.8%) | 259 (50.3%) | <0.001 | 1,407 (71.7%) | 1,407 (68.8%) | 0.050 | <0.001 | 0.101 |
| Complementary feeding age | 7224 | 5,666 (78.4%) | 5,370 (80.0%) | 296 (57.5%) | <0.001 | 1,552 (79.1%) | 1,650 (80.7%) | 0.215 | <0.001 | 0.215 |
| Walking red flag at 2 yrs | 7224 | 6,010 (83.2%) | 5,619 (83.8%) | 391 (75.9%) | <0.001 | 1,635 (83.3%) | 1,736 (84.9%) | 0.180 | <0.001 | 0.215 |
| Composite literacy score (0-4) | 7224 | median 3 [3-4]; mean 3.19 (SD 0.91) | 3.24 (0.87) | 2.60 (1.11) | <0.001 | 3.26 (0.87) | 3.24 (0.88) | 0.452 | - | - |

**Multimedia Appendix 13.**
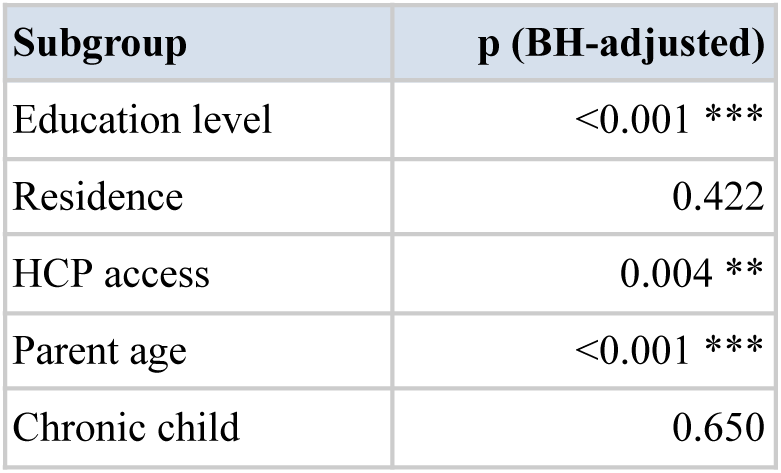
Child-health literacy score, by sociodemographic subgroup. Questionnaire 2 (n = 7,224). Composite literacy score (0 to 4). Tests: Kruskal-Wallis for ordinal subgroups, Mann-Whitney for binary chronic-child. p (BH-adjusted) : BH-FDR across 5 tests.

| Subgroup | p (BH-adjusted) |
| --- | --- |
| Education level | <0.001 *** |
| Residence | 0.422 |
| HCP access | 0.004 ** |
| Parent age | <0.001 *** |
| Chronic child | 0.650 |

**Multimedia Appendix 14.** AI use patterns, full detail. Questionnaire 2 (n=7,224; AI users n=3,919). Frequency of AI use, query topics among users, twelve-month trend among respondents who reported a direction of change (n=3,089), and perceived useful situations. Descriptive only; subgroup associations with AI use are reported in Multimedia *Appendix 5* and *Figure 2*.

| Item | n (%) of denominator | Denominator |
| --- | --- | --- |
| **AI use: frequency** |  |  |
| Never used | 3,305 (45.8%) | all respondents (n=7,224) |
| Once or twice (curiosity) | 2,185 (30.2%) | all respondents (n=7,224) |
| Several times per month | 928 (12.8%) | all respondents (n=7,224) |
| Several times per week | 806 (11.2%) | all respondents (n=7,224) |
| Any use (binary) | 3,919 (54.2%) | all respondents (n=7,224) |
| **AI use: query topics** |  |  |
| Symptoms & illnesses | 2,349 (59.9%) | AI users (n=3,919) |
| Activity / play ideas | 1,866 (47.6%) | AI users (n=3,919) |
| Sleep (troubles, methods) | 1,645 (42.0%) | AI users (n=3,919) |
| Feeding & nutrition | 1,526 (38.9%) | AI users (n=3,919) |
| Behavior & emotions | 1,435 (36.6%) | AI users (n=3,919) |
| Everyday practical questions | 1,384 (35.3%) | AI users (n=3,919) |
| Child development | 1,402 (35.8%) | AI users (n=3,919) |
| Vaccination | 451 (11.5%) | AI users (n=3,919) |
| **AI use: 12-month evolution** |  |  |
| Greatly decreased | 101 (3.3%) | reporters (n=3,089; excludes 'was not using one year ago') |
| Slightly decreased | 91 (2.9%) | reporters (n=3,089; excludes 'was not using one year ago') |
| Unchanged | 788 (25.5%) | reporters (n=3,089; excludes 'was not using one year ago') |
| Slightly increased | 954 (30.9%) | reporters (n=3,089; excludes 'was not using one year ago') |
| Greatly increased | 1,155 (37.4%) | reporters (n=3,089; excludes 'was not using one year ago') |
| **Perceived useful situations** |  |  |
| Everyday questions not needing a doctor | 3,247 (44.9%) | all respondents (n=7,224) |
| Reassurance on minor worries | 2,765 (38.3%) | all respondents (n=7,224) |
| At night (emergency triage) | 2,138 (29.6%) | all respondents (n=7,224) |
| First orientation before appointment | 1,982 (27.4%) | all respondents (n=7,224) |
| No situation feels useful | 1,820 (25.2%) | all respondents (n=7,224) |
| Verifying info found elsewhere | 1,399 (19.4%) | all respondents (n=7,224) |
| Tracking child development | 956 (13.2%) | all respondents (n=7,224) |
| **Number of useful situations (per parent)** | median 1 [1-3]; mean 1.73 (SD 1.58) | all respondents (n=7,224) |

**Multimedia Appendix 15.**
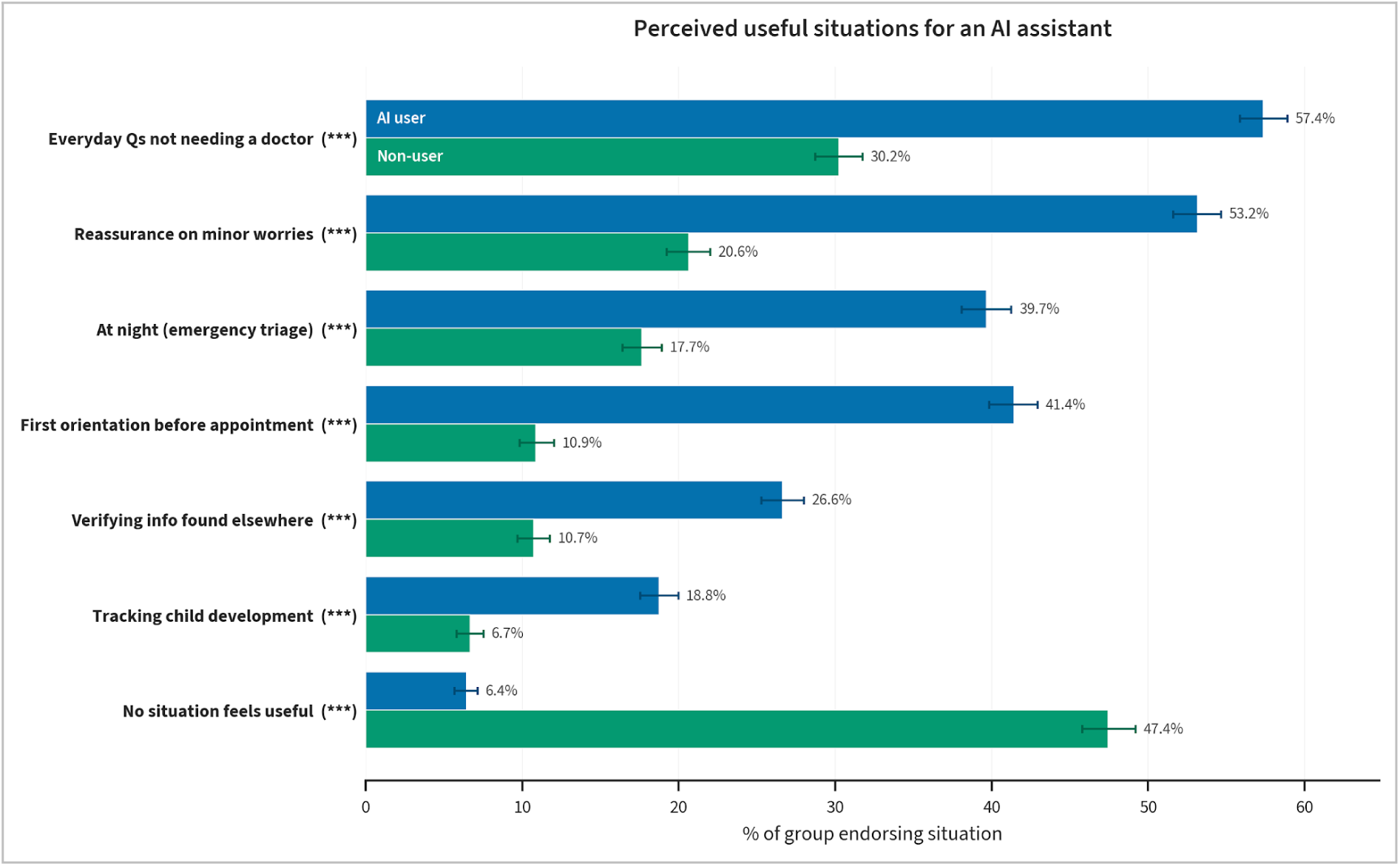
Perceived useful situations for an AI assistant, AI users vs non-users. Questionnaire 2 (AI users n=3,919; non-users n=3,305). Percentage of each group endorsing each situation. Error bars are bootstrap 95% confidence intervals. Comparison by chi-square test (users vs non-users). Significance (Benjamini-Hochberg-adjusted): *P<.05, **P<.01, ***P<.001.

**Multimedia Appendix 16.** Barriers, trust enablers, impact on consultations, and disclosure to AI. Questionnaire 2 (n = 7,224). Barriers to AI use, conditions for trusting an AI tool, perceived effect on the frequency of professional consultations (AI users who answered, n = 3,689), and disclosure to AI of concerns shared with neither a professional nor the personal network. Comparisons across the three user categories by chi-square with Fisher fallback, with BH correction within each family; the consultation-impact section is descriptive only. Raw and Benjamini-Hochberg-adjusted p-values (p adj.) across ten tests are reported.

| Item | Overall n (%) | Non-user | Occasional | Regular | p | p (BH-adjusted) | Denominator |
| --- | --- | --- | --- | --- | --- | --- | --- |
| **Perceived barriers to AI use** |  |  |  |  |  |  |  |
| Prefer talking to a professional | 4,037 (55.9%) | 2,146 (64.9%) | 1,228 (56.2%) | 663 (38.2%) | <0.001 | <0.001 | all (n=7,224) |
| Concerns about reliability | 3,517 (48.7%) | 1,641 (49.7%) | 1,174 (53.7%) | 702 (40.5%) | <0.001 | <0.001 | all (n=7,224) |
| Nothing holds me back | 1,373 (19.0%) | 275 (8.3%) | 387 (17.7%) | 711 (41.0%) | <0.001 | <0.001 | all (n=7,224) |
| Personal data privacy concerns | 1,048 (14.5%) | 494 (14.9%) | 339 (15.5%) | 215 (12.4%) | 0.014 | 0.014 | all (n=7,224) |
| Not comfortable with these tools | 476 (6.6%) | 372 (11.3%) | 77 (3.5%) | 27 (1.6%) | <0.001 | <0.001 | all (n=7,224) |
| **Trust enablers** |  |  |  |  |  |  |  |
| Supervised by physicians / pediatricians | 5,537 (76.6%) | 2,347 (71.0%) | 1,762 (80.6%) | 1,428 (82.4%) | <0.001 | <0.001 | all (n=7,224) |
| Certified by an official authority (HAS, Min. Santé) | 4,181 (57.9%) | 1,787 (54.1%) | 1,323 (60.5%) | 1,071 (61.8%) | <0.001 | <0.001 | all (n=7,224) |
| Recommended by my doctor | 2,467 (34.2%) | 1,167 (35.3%) | 765 (35.0%) | 535 (30.9%) | 0.004 | 0.004 | all (n=7,224) |
| Personal data protected | 2,272 (31.5%) | 946 (28.6%) | 717 (32.8%) | 609 (35.1%) | <0.001 | <0.001 | all (n=7,224) |
| Transparent sources of information | 2,205 (30.5%) | 836 (25.3%) | 714 (32.7%) | 655 (37.8%) | <0.001 | <0.001 | all (n=7,224) |
| Free of charge | 2,017 (27.9%) | 763 (23.1%) | 615 (28.1%) | 639 (36.9%) | <0.001 | <0.001 | all (n=7,224) |
| Tested / approved by other parents | 705 (9.8%) | 274 (8.3%) | 208 (9.5%) | 223 (12.9%) | <0.001 | <0.001 | all (n=7,224) |
| **Impact on consultations (AI users only)** |  |  |  |  |  |  |  |
| More often (AI flags issues I would miss) | 274 (7.4%) |  |  |  |  |  | AI users who answered (n=3,689) |
| As often as before | 2,466 (66.8%) |  |  |  |  |  | AI users who answered (n=3,689) |
| Less often (AI reassures me enough) | 253 (6.9%) |  |  |  |  |  | AI users who answered (n=3,689) |
| Differently: I come better prepared | 696 (18.9%) |  |  |  |  |  | AI users who answered (n=3,689) |
| **Disclosure to AI (concerns not shared with a professional or network)** |  |  |  |  |  |  |  |
| Any disclosure to AI | 2,563 (35.5%) | 508 (15.4%) | 943 (43.2%) | 1,112 (64.1%) | <0.001 | - | all (n=7,224) |
| Personal well-being (fatigue / anxiety / mood) | 1,821 (25.2%) | 291 (8.8%) | 648 (29.7%) | 882 (50.9%) | <0.001 | <0.001 | all (n=7,224) |
| Other topics | 850 (11.8%) | 219 (6.6%) | 288 (13.2%) | 343 (19.8%) | <0.001 | <0.001 | all (n=7,224) |
| Doubts about my parenting role | 791 (10.9%) | 71 (2.1%) | 255 (11.7%) | 465 (26.8%) | <0.001 | <0.001 | all (n=7,224) |
| Questions I did not dare ask a doctor | 571 (7.9%) | 48 (1.5%) | 205 (9.4%) | 318 (18.3%) | <0.001 | <0.001 | all (n=7,224) |

